# A new, simple method of describing the COVID-19 trajectory and dynamics in any country based on Johnson Cumulative Distribution Function fitting

**DOI:** 10.1101/2020.12.05.20244178

**Authors:** Adam M. Ćmiel, Bogdan Ćmiel

## Abstract

A simple method is described to study and compare COVID-19 infection dynamics between countries, based on curve fitting to publicly shared data of confirmed COVID-19 infections in them. The method was tested using data from 80 countries from 6 continents. We found that Johnson Cumulative Distribution Functions (CDF) were extremely well fitted to the data (R^2^>0.99) and that Johnson CDFs were much better fitted to the data at their tails than either the commonly used Normal or Lognormal CDFs. Fitted Johnson CDFs can be used to obtain basic parameters of the infection wave, such as the percentage of the population infected during an infection wave, the days of the start, peak and end of the infection wave, as well as the durations of the infection wave of the wave’s increase and decrease. These parameters can be easily interpreted biologically and used both for describing the infection wave dynamics and in further statistical analysis. The usefulness of the parameters obtained was analysed with respect to the relation between the Gross Domestic Product (GDP) per capita and the population density, and the percentage of the population infected during an infection wave, the starting day and the duration of the infection wave in the 80 countries. We found that all the above parameters were significantly dependent on the GDP per capita, but only the percentage of the population infected was significantly dependent on the population density in these countries. If used with caution, this method has a limited ability to predict the future trajectory and parameters of an ongoing infection wave.

## Introduction

A highly contagious disease, COVID-19 is caused by the SARS-CoV-2 coronavirus. This virus was first detected in Wuhan (central China) in December 2019, but as early as mid-January 2020, the virus had quickly spread throughout China. On 13 January 2020, the first case outside China was confirmed and on 24 January, the first case in Europe was reported. In the second half of February 2020, outbreaks with hundreds of cases erupted in South Korea, Italy and Iran (Skórka et al., 2020), and COVID-19 was declared a pandemic by the World Health Organization on 11 March 2020 (Ducharme, 2020). To date, over 64 million infections and almost 1.5 million deaths have been reported globally (WHO, 2020).

Since the very beginning of the pandemic, many models have been proposed to understand the outbreak dynamics of COVID-19 (e.g. IHME, 2020; UGSDSC, 2020; LANL, 2020; Ferguson et al., 2020; Kissler et al., 2020; Aleta et al.; Hellewell et al., 2020) and have been used by policymakers (e.g. the US Government) for allocating resources or planning interventions. Some of them, such as the early IHME model, have received a fair amount of criticism (Jewell et al., 2020). COVID-19 modelling studies generally follow one of two general approaches – forecasting models and mechanistic models – although there are also hybrid approaches (Holmdahl and Buckee, 2020). Forecasting models are often statistical in nature, fitting a line or a curve to data and extrapolating from there, without incorporating the process that produces the pattern (Holmdahl and Buckee, 2020), while mechanistic models simulate the outbreak through interacting disease mechanisms by using local nonlinear population dynamics and the global mixing of populations (Hethcote, 2000). Purely statistical models are reliable only within a short time window and may be useful for making rapid short-term recommendations, whereas mechanistic modelling can be useful for exploring how the course of the pandemic might change under various assumptions and political interventions (Kuhl, 2020).

Since its onset, the COVID-19 pandemic has generated a huge amount of data and is probably the best documented disease in history. New cases, active cases, deaths and numbers of tests performed are usually published daily by official sources (e.g. governments), gathered and publicly shared as freely accessible datasets (e.g. Hasell et al., 2020). This offers researchers an opportunity to focus on analysing the pandemic and its dynamics in fields other than epidemiology. Although the above-mentioned models provide many pandemic parameters for predicting different scenarios of future infections, the probability and duration of future pandemic peaks, which is extremely useful for policymakers in planning interventions, they may not be very useful in fields other than epidemiology. An urgent need thus arose to develop methods of describing the trajectories of pandemic waves. Such methods should be simple to apply and should provide parameters describing the trajectory and dynamics of the epidemic that are easy to interpret and employ in further statistical analyses by researchers in other fields, like sociology, biology and ecology, which can deepen our understanding of the COVID-19 pandemic.

The aim of this paper is to present a new and simple method based on curve fitting to the reported data on confirmed cases of infection, and to study and compare the infection dynamics between countries (or regions). This method, based on Johnson Cumulative Distribution Function (CDF) fitting, was tested using data from 80 countries from 6 continents (Africa, Asia, Europe, Oceania, as well as North and South America). Also, Johnson CDFs were used to calculate basic parameters of the infection wave dynamics, such as the percentage of the population infected during the infection wave, the days of the start, peak and end of the infection wave, the duration of the infection wave, and the durations of the wave’s increase and decrease. These parameters are easy to interpret and can be used in further statistical analyses of epidemic dynamics. This is exemplified by the influences of Global Domestic Product (GDP) per capita and of population density on the percentage of infections and the first day and duration of the first infection wave in the countries concerned. The method presented here and the techniques employed are both straightforward and well known; they illustrate how simple techniques can be used to solve otherwise difficult problems, such as describing an epidemic wave.

## Materials and methods

The data used in this study were obtained from the Our World in Data COVID-19 dataset (Hasell et al., 2020) from December 2019 to 19 November 2020. The method was tested on 80 countries from 6 regions: 1) Africa (Democratic Republic of Congo, Egypt, Ethiopia, Kenya, Morocco, Nigeria, Somalia, South Africa, South Sudan, Sudan and Zimbabwe); 2) Asia (Afghanistan, Bangladesh, Cambodia, China, India, Indonesia, Iran, Iraq, Israel, Japan, Lebanon, Myanmar, Pakistan, Philippines, Saudi Arabia, Singapore, South Korea, Sri Lanka, Syria, Taiwan, Thailand, Turkey, Vietnam); 3) Europe (Austria, Belgium, Bosnia and Herzegovina, Bulgaria, Croatia, Cyprus, Czechia, Finland, France, Germany, Greece, Hungary, Ireland, Italy, Netherlands, North Macedonia, Norway, Poland, Portugal, Romania, Russia, Serbia, Slovakia, Slovenia, Spain, Sweden, Switzerland, Ukraine, United Kingdom); 4) North America (Canada, Jamaica, Mexico, United States of America); 5) Oceania (Australia, Fiji, New Zealand, Papua New Guinea); and 6) South America (Argentina, Bolivia, Brazil, Chile, Colombia, Paraguay, Peru, Uruguay, Venezuela).

In order to make the data comparable between countries, the number of infections on each day of the pandemic for each country were standardized – they are presented here as a percentage of the population of a given country infected, i.e. the number of confirmed infections in a given country/country population*100%. Also, a five-day moving average was calculated using the percentage of infections to smooth the data and to minimize the effect of a smaller number of tests performed and a smaller number of confirmed infections during some short periods like weekends. This makes the loss function more regular, i.e. it has less relative extrema, and so it is easier to find the global extremum. Nevertheless, all the presented R^2^ for the Johnson CDFs obtained were calculated using raw (not smoothed) data.

### Fitting Johnson CDF by moments

Johnson (1949) described a system of frequency curves that represents transformations of the standard normal curve (detailed description in Hahn and Shapiro, 1967). Applying these transformations to a standard normal variable allows a unique distribution to be derived for whatever combination of mean, standard deviation, skewness and kurtosis occurs for a given set of observed data. The standard method of fitting Johnson curves is to use four coefficients defining a Johnson distribution: two shape coefficients (*γ, δ*), as well as a location (*ξ*) and a scale (*λ*) coefficient:

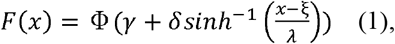

where Φ is the cumulative distribution function of the standard normal distribution. This method is not intuitive, however, as it is difficult to set up starting points from the data in order to perform numerical fitting. Thus, we selected an alternative method for fitting Johnson curves, using the first four moments of an empirical distribution (mean, variance, skewness and kurtosis; detailed descriptions in Hahn and Shapiro (1967) and Hill et al. (1976)). All the statistical fits in the paper were performed using the Levenberg-Marquardt algorithm (Moré, 1978) to solve the corresponding non-linear least square optimization problem. The convergence criterion was set at 1.0E^-10^.

### Fitting Johnson CDFs to epidemic waves

There is no strict definition of what is or is not an epidemic wave or phase. The intuitive definition of a pandemic wave traces the development of an epidemic over time and/or space. During an epidemic, the number of new cases of infection increases (often rapidly) to a peak and then falls (usually more gradually) until the epidemic wave is over.

The epidemic’s dynamics may differ greatly between countries. Since the beginning of the pandemic, there has been only one epidemic wave in some countries (e.g. Afghanistan, Argentina), while in others two have occurred (e.g. Australia); in yet others, even more have taken place, which may have overlapped and interfered with each other, as in Croatia, where there were four overlapping and interfering waves. The authorities in many countries, moreover, have imposed lockdowns of varying severity in order to slow down or “flatten” the infection curve. Hence, the epidemic waves may not follow Farr’s law (which states that epidemics tend to rise and fall in a roughly symmetrical pattern or bell-shaped curve) and may be asymmetrical.

The basic assumption is that each epidemic wave *W* in a given country can be described by a five-parameter scaled Johnson CDF: scale parameter (*s*), and the above-mentioned moments– expected value (mean, *E*), variance (*V*), skewness (*S*) and kurtosis (*K*)

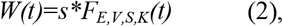

where *t* is the time measured since the day of the beginning of the pandemic, and the function *F*_*E,V,S,K*_ is the Johnson CDF with parameters *γ, δ, ξ, λ* assuming the mean, variance, skewness and kurtosis to be equal to *E,V,S,K*, respectively (see Hahn and Shapiro,1967; Hill et al.,1976). Parameters *S* and *K* were expected to improve the curve fit at the tails of the epidemic wave if it was not symmetrical or heavy tailed.

### Obtaining basic epidemic wave parameters and their biological interpretation

Once the Johnson CDFs were fitted to each pandemic wave in a given country, the basic parameters for obtaining the wave dynamics, i.e. 2.5% quantile (*Q*_*2*.*5%*_), 50% quantile (median; *Q*_*50%*_), 97.5% quantile (*Q*_*97*.*5%*_), could be calculated:

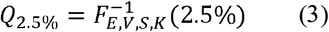

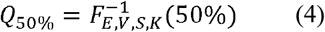

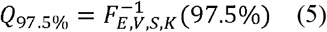

The disadvantage of fitting the Johnson curve by its moments is that it is not possible to determine its mode analytically. Thus, the mode of each Johnson CDF was determined numerically:

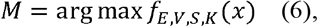

where *f*_*E,V,S,K*_ is the Johnson Probability Density Function (PDF).

These parameters have an intuitive biological interpretation (Fig. 1). The scale parameter *s* indicates the total percentage of infections during a given epidemic wave (*P*_*inf*_), *Q*_*2*.*5%*_ indicates the day when the infection wave started, while *Q*_*97*.*5%*_ indicates its end. The median (*Q*_*50%*_) indicates the day when half the total percentage of infections during a given wave was reached. Finally, the mode (*M*) indicates the day when the peak occurred. In addition, one can easily obtain the wave duration (*T*)

**Figure 1.**
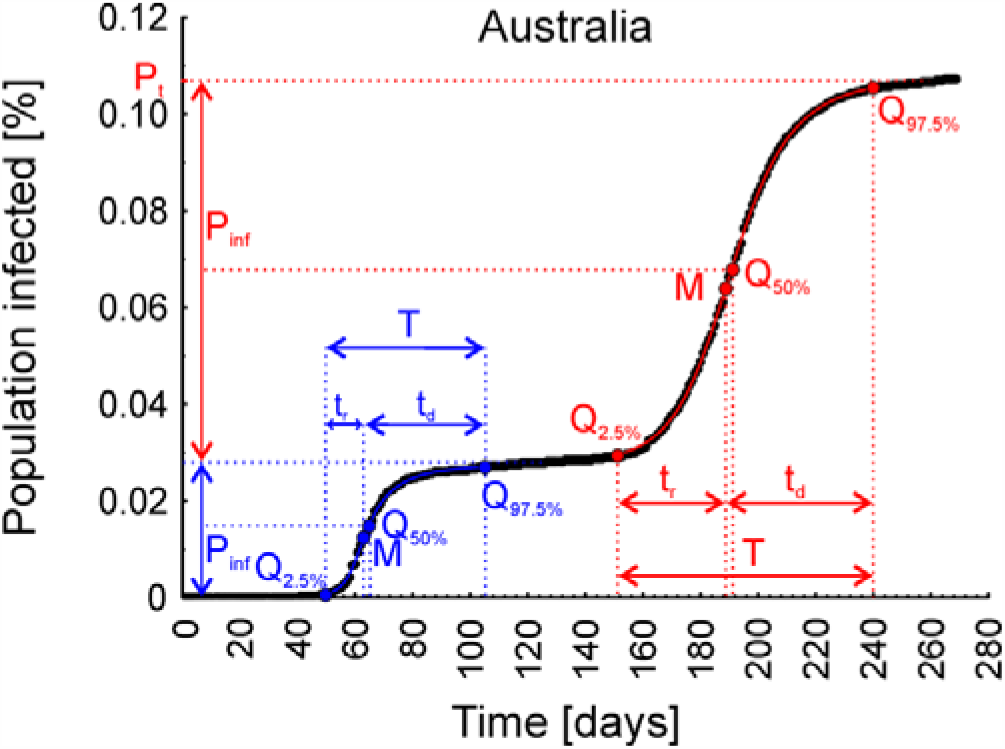
Graphical presentation of the interpretation of the parameters obtained from Johnson Cumulative Distribution Function fitting, describing the dynamics of the two infection waves observed in Australia. *P*_*inf*_ – the total percentage of infections in a given infection wave, *Q*_*2*.*5%*_ – the day when the infection wave started, *Q*_*97*.*5%*_ – the day when the infection wave ended, *Q*_*50%*_ – the day when half of the total percentage of infections during a given wave was reached, *M* – the day when the infection wave peaked, *T* – the duration of the wave, *t*_*i*_ – the duration of the wave increase, *t*_*d*_ – the duration of the wave decrease, *P*_*t*_ – the total percentage of the population infected after two infection waves.

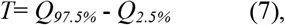

the duration of wave increase (*t*_*i*_)

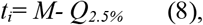

and the duration of the wave decrease (*t*_*d*_)

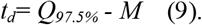

Also, the parameter measuring the asymmetry of the infection wave (*A*) is easily obtained as the ratio

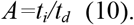

All of the above-mentioned parameters can be easily used in further statistical analyses, as exemplified by 1) the relationship between Gross Domestic Product (GDP) per capita and basic parameters describing the dynamics of the first wave of infections: *M, T* and *P*_*inf*_, and 2) the relation between population density and basic parameters describing the dynamics of the first wave of infections: *M, T* and *P*_*inf*_. Only the first wave of infections in each country has been taken into account here, because in some countries, second (and subsequent) waves did not take place and would have to be excluded from the analysis.

### Comparing curves: Johnson vs Normal and Lognormal CDFs

The differences between the Johnson, Normal and Lognormal CDFs were highlighted on the basis of data from Afghanistan, where only one epidemic wave took place, by comparing parameters R^2^, *P*_*inf*_, *Q*_*2*.*5%*_, *M* and *Q*_*97*.*5%*_. Both the 2.5% and the 97.5% quantiles for normal and lognormal distributions were obtained using inverse Normal and inverse Lognormal PDFs, respectively.

### Fitting Johnson’s CDF to the ongoing wave and forecast possibilities

Fitting Johnson’s curve to the ongoing wave yields parameters that can also be interpreted as a forecast of the future shape and dynamics of the infection wave. In such a case, *P*_*inf*_, *M* and *Q*_*97*.*5%*_ indicate the predicted percentage of infections, the predicted day of the peak and the predicted day of the end of the ongoing wave, respectively, which can also be used to calculate the predicted times of the increase, decrease and duration of the ongoing infection wave. Because this method is intended to describe the infection’s dynamics rather than to predict its ultimate outcome, the accuracy of the forecast is evaluated only on the basis of data from the first wave of infection recorded in the United Kingdom (see Supplementary Materials).

### Examples of application

#### The relationships between Gross Domestic Product (GDP) per capita, and population density and the dynamics of the first wave of COVID-19 infections

The data on the GDP per capita and population density in the 80 countries analysed here were obtained from the Our World in Data COVID-19 dataset (Hasell et al., 2020).

The relationship between GDP per capita, and population density and the basic parameters describing the dynamics of the first wave of infections (*M, T* and *P*_*inf*_), obtained using the presented method of Johnson CDF fitting, was tested using the quantile dependence function method, described in detail in Ćmiel and Ledwina (2020).This method was designed for measuring, visualizing the dependence structure, and testing the independence of two random variables. It exploits a recently introduced local dependence measure (quantile dependence function *q*), which gives a detailed picture of the underlying dependence structure and provides a means by which the local association structure can be minutely examined at different quantile levels (Ćmiel and Ledwina 2020).

## Results

Fig. 2 illustrates examples of Johnson curves fitted to the data from countries where one ongoing infection wave (Argentina), one infection wave (Afghanistan), two infection waves (Australia) and four overlapping and interfering infection waves (Croatia) occurred. Fig. 3A shows Johnson CDFs fitted to the four waves of infections observed in Croatia, showing where the waves overlapped and interfered.

**Fig. 2.**
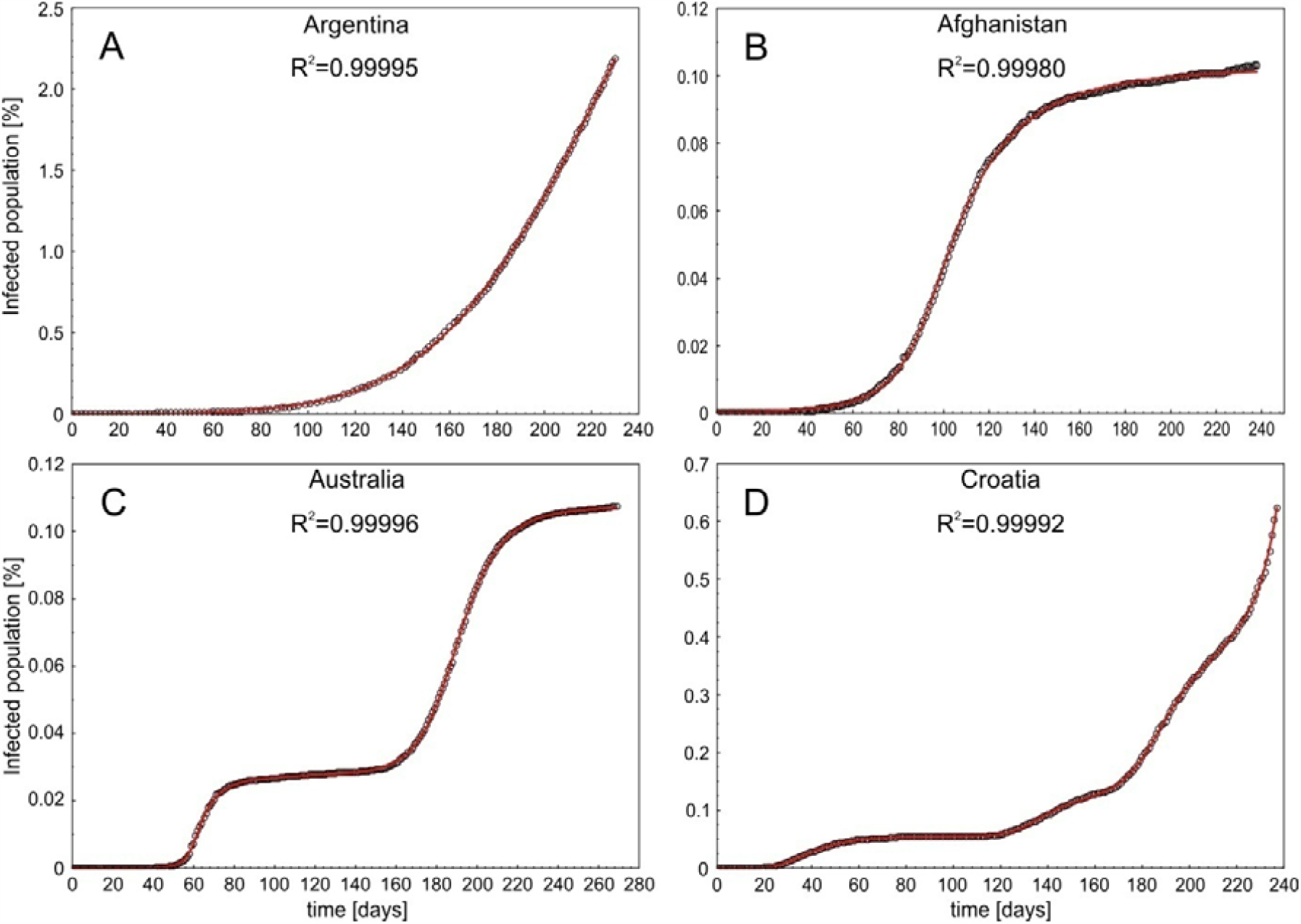
Examples of fitted distributions in four scenarios of COVID-19 infection dynamics. A – one ongoing infection wave (before the peak), B – one full wave, C – two waves and D – four overlapping and interfering waves. Open circles – raw data, red lines – fitted Johnson Cumulative Distribution Functions.

**Fig. 3.**
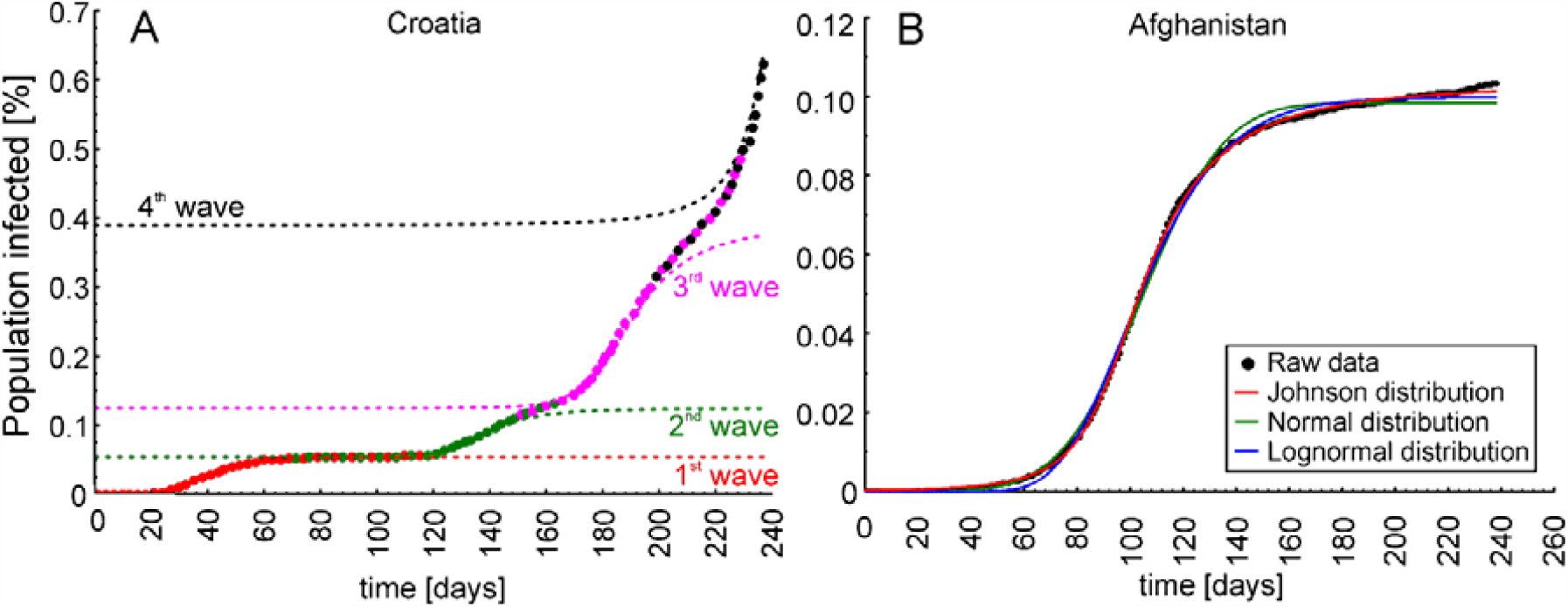
A – Trajectory of four Johnson Cumulative Distribution Functions fitted to the four waves of infections reported in Croatia, showing the areas where the waves overlapped and interfered. B – the differences between Johnson (red line), Normal (green line) and Lognormal Cumulative Distribution (blue line) Functions fitted to the raw data from Afghanistan (black dots).

Johnson CDF fitting, tested using data from 80 different countries, showed that all the curves were extremely well fitted: the lowest R^2^ was 0.995 (Fiji), the highest R^2^ was 0.99997 (Iraq), and the mean and median R^2^ were 0.9995 and 0.9997, respectively. The functions fitted with R^2^ and the COVID-19 trajectory plots with fitted functions for each country are illustrated in the Supplementary Materials (Table S1; Figures S1-S6).

Fitting Johnson, Normal and Lognormal distribution curves to the single wave of infection that took place in Afghanistan showed that the Johnson CDF (R^2^=0.9998) was the best fitted, whereas the Normal (R^2^=0.9980) and Lognormal (R^2^=0.9989) distributions were not so well fitted, mainly at the tails of the infection wave (Fig. 3B). The parameters *Q*_*2*.*5%*_, *M* and *Q*_*97*.*5%*_ obtained for the infection wave in Afghanistan using Johnson CDF fitting were 59, 100 and 209 respectively, whereas the same parameters obtained using Normal CDF fitting and Lognormal CDF fitting were 57, 105 and 152, and 65, 98 and 167, respectively. The percentages of the population infected during the infection wave obtained using the scale parameters (*s*) of the fitted Johnson, Normal and Lognormal distributions were 0.1028%, 0.0984% and 0.0997% respectively.

Seventeen (21.3%) of the 80 countries analysed were described by fitting one infection wave, while 35 (43.8%), 24 (30%) and 4 (5%) were described by fitting two, three and four infection waves, respectively (Table S1).

The basic statistics for the skewness parameters of the Johnson CDFs fitted to the first pandemic waves in the 80 counties showed that in the majority of them, the first wave of infection was skewed (median *S*=1.5; minimum *S*=0; maximum *S*=141.5). The first wave of infection was symmetrical in 16 countries (20%; *A<*1.05). Also, the basic statistics for parameter *A* showed that the duration of wave decrease was longer than that of wave increase (mean *A*= 4.7; median *A*=2.9; minimum *A*=1.0; maximum *A*=22.4).

Analysis of the associations between GDP per capita and parameters *M, T* and *P*_*inf*_ showed that the percentage of confirmed infections during the first epidemic wave in the 80 countries was dependent on the GDP per capita (p=0.0147; Fig 4A), the time of the peak (*M*; p=0.0002; Fig. 4B) and the duration of the first epidemic wave (*T*; p=0.0087; Fig. 4C). The relation between the percentage of infections and GDP per capita tended towards a global positive dependence (Fig. 4A), which means that the higher the GDP per capita, the greater the percentage of infections during the first epidemic wave. The relation between the time of the peak and GDP per capita showed a local negative dependence for countries where the peak occurred late (above median; Fig. 4B). This means that the very early occurrence of a peak was not really correlated with GDP per capita; but when the peak did not occur early, then the higher the GDP per capita, the earlier the peak occurred. A similar situation prevailed for the relation between the duration of the infection wave and GDP per capita (Fig. 4C), i.e. a very short duration of the first epidemic wave was not really correlated with GDP per capita, but in the case when the first epidemic wave was not of short duration, then the higher the GDP per capita, the shorter the first epidemic wave.

**Fig. 4.**
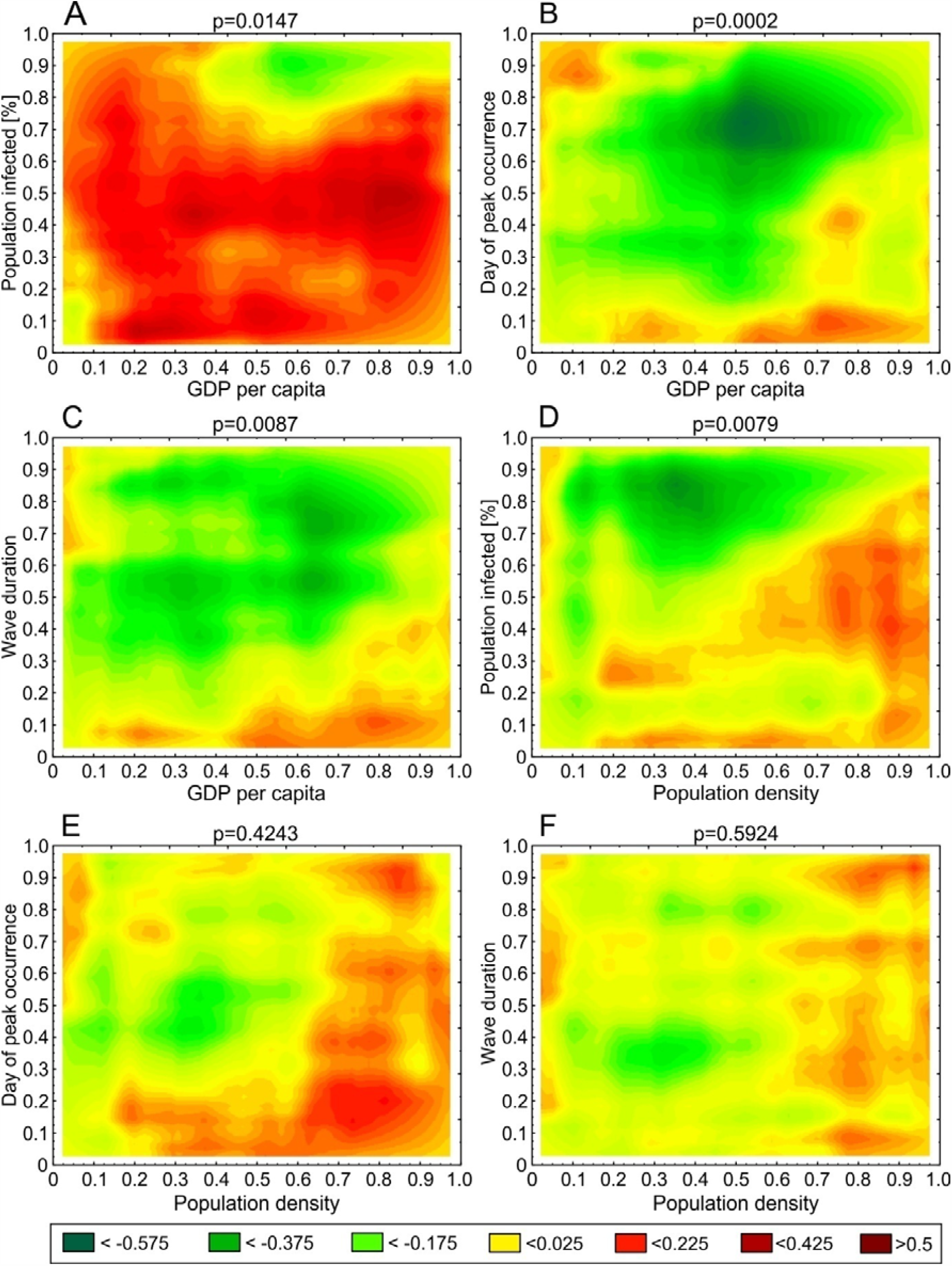
Heat maps showing the local association structure between variables at different quantile levels obtained using the quantile dependence function *q*.

Analysis of the associations between population density and parameters *M, T* and *P*_*inf*_ showed that the percentage of infections during the first epidemic wave in the 80 countries depended on population density (p=0.0079; Fig 4D), whereas the day of the peak and the duration of the first epidemic wave did not (*T*: p=0.4243; Fig. 4E; *M*: p=0.5924; Fig. 4F).The relation between the percentage of infections and population density showed a local negative dependence (Fig. 4D), e.g. in cases when the population density was not very high but the percentage of infections was quite high. In such cases, the greater the population density, the smaller the percentage of infections.

## Discussion

The method presented in this paper gives an indication of the spread of COVID-19 in any particular country that publicizes daily numbers of infected cases. Both this method and the techniques employed are all straightforward, well known and easy to use, since Johnson CDF fitting is available in many statistical/calculus packages, e.g. R, Statistica, MATLAB, MS Excel. By using an alternative method of fitting using moments instead of shape, location and scale parameters, it is easier to set starting points for numerical fitting, e.g. by visually analysing the scatter plot of the number of infected cases in time. The curves are extremely well fitted: this is exemplified by the data from 80 different countries on 6 continents. Also, the parameters are easy to interpret and ready to use in further analyses, such as finding associations between them and other variables that may be associated with COVID-19 dynamics, i.e. GDP per capita, population density.

Some researchers have used curve-fitting with a normal distribution to respond to a real time request, applying it to COVID-19 in Wuhan (Tomie, 2020), since it was known that flu epidemics follow a normal distribution, whereas other researchers noticed the COVID-19 profile had a characteristically asymmetric tail and applied Lognormal distribution curve fitting (Nishimoto and Inoue, 2020). Our results show that the first wave of infections was highly skewed in 79% of the countries analysed. This suggests that unlike flu epidemics, the COVID-19 epidemic is not following a normal distribution and should not be modelled in this manner. In such a case, log-normal distribution fitting appears better; but as the example of Afghanistan shows, the apparent differences in R^2^ between the Johnson, Normal and Lognormal CDFs are small, but are in fact ca one order of magnitude in favour of the Johnson CDF. Moreover, one can see that both the Normal and Lognormal CDFs are less well fitted at the tails of the infection wave than the Johnson CDF (Fig. 3B), and both reveal a smaller number of infections than was actually recorded (raw data) and lower than that obtained using Johnson’s CDF. Moreover, the fitted Lognormal curve starts to increase later than the Normal and Johnson distribution curves, which would in consequence lead to an incorrect estimate of the beginning of the wave (11 days later than when obtained using Johnson’s distribution), whereas the Normal distribution is far worse fitted at the right tail than the Johnson and Lognormal distributions, because the infection wave in Afghanistan was not symmetrical. Apart from the fact that using the Normal distribution would preclude any estimate of the true duration of the wave decrease (by definition equal to the time of the wave increase), it also leads to a much lower estimate of the day when the wave of infections ends (57 days earlier than estimated using the Johnson distribution), which is caused by the “too fast” flattening of the Normal CDF (Fig. 3B). The extremely high R^2^ values obtained for the 80 countries (see Supplementary Materials) suggest that the Johnson curve class is flexible enough to almost perfectly follow the course of the epidemic in these countries. This is because both skewness and kurtosis are estimated during the Johnson curve fitting procedure, whereas the shapes of other commonly used curves (Normal, Lognormal, Weibull) are more or less imposed. This result also suggests that the Johnson distribution should be the preferred curve-fitting approach for COVID-19 data.

The curve fitting method presented here was designed primarily to obtain easily interpretable parameters describing past trajectories of COVID-19 infections, but the parameters describing the current wave of infection, especially in its early stages (before the peak), can be interpreted as a forecast of the future course of the pandemic. In such a case, however, extreme caution is advisable (see Jewell et al., 2020). This method is a purely statistical model that does not incorporate the process producing the pattern of the number of infections and does not account for parameters governing transmission, disease and immunity. In addition, curve fitting techniques cannot predict the occurrence of future peaks. Thus, for long-term forecasts and modelling future scenarios of the pandemic, it is recommended to use more reliable methods, such as those based on SEIR models. Nevertheless, some short-term predictions can be made using the Johnson CDF method, which may be useful to policymakers for planning rapid, short-term interventions. One must bear in mind, however, the method’s limitations (see above), as well as those resulting from the data collection and reporting, which are discussed later in this section.

The results obtained from this application of parameters describing COVID-19 dynamics have shown that the higher the GDP per capita, the higher the percentage of the population infected. This is quite an unexpected result, but consistent with a very recent report by Liu et al. (2020), who found a positive correlation between the human development index (HDI) and the risk of infection and death from COVID-19 in Italy. Other results have shown that, excluding countries where the infection wave peaked very early and its duration was short, the higher the GDP per capita, the earlier the peak and the shorter the first epidemic wave. This result, in turn, is similar to that reported in another very recent paper, in which the date of the first COVID-19 cases was shown to co-vary positively with GDP across countries, most probably because of their closer involvement in global tourism and transport (Jankowiak et al., 2020). Another example showed that the higher the population density, the lower the percentage of a population infected during the first wave of infections. This, too, seems unexpected; but this negative dependence results from the fact that infections are presented as a percentage, which does not scale proportionally with population density. A further possible explanation is that in countries with a high population density, like China and Singapore, very strict (full) lockdowns were immediately applied (China, Kretschmer and Yang, 2020; Singapore, Cheong, 2020), which could have resulted in fewer people being infected than in countries with a lower population density, where lockdowns were only partial, if imposed at all. Moreover, some researchers report a positive correlation between population density and the number of infections and related mortality, e.g. in India (Bhadra et al., 2020),whereas others provide no evidence that population density is linked with COVID-19 cases and deaths, e.g. the USA (Carozzi et al., 2020). Nevertheless, these examples demonstrate the usefulness of our method. The very recent papers by Liu et al. (2020) and Jankowiak et al. (2020) have also shown that the amount of research on COVID-19, other than purely epidemiological modelling of future pandemic scenarios, is increasing. This indicates that simple methods of obtaining parameters describing infection waves, such as are presented in this paper, may be very useful and can help to deepen our understanding of the COVID-19 pandemic.

The last, but not least important issue which has to be addressed is a key limitation in understanding the COVID-19 pandemic, namely, that the true number of infections is not known and the only infections known are those confirmed by tests. Moreover, testing strategies differ between countries: in some countries only symptomatic cases are tested, while in others mass testing is carried out. Moreover, most COVID-19 cases are asymptomatic and remain unreported (Peirlinck et al., 2020). Consequently, mortality data are generally considered more reliable than the testing-dependent confirmed case counts that are used in COVID-19 epidemic modelling (e.g. Chikobvu and Sigauke, 2020). However, some countries only report COVID-19 deaths occurring in hospitals, whereas others report COVID-19 deaths when a test has confirmed the infection (this makes the number of mortalities testing-dependent as well). On the other hand, if laboratory diagnosis is not required, as in the U.K. (UK Guidance), it is possible that deaths due to other diseases with COVID-19-like symptoms are being reported as COVID-19 deaths. It may also be difficult to specify the cause of death in cases where patients had other diseases, e.g. an advanced stage of cancer, co-existing with COVID-19. Taking all of the above into account, it is very likely that the real number of deaths is also higher than the reported number of deaths, something that has been noticed in some countries, e.g. Italy (Foresti, 2020; Stancati and Sylvers, 2020) and China (Long et al., 2020). It is may well be that the numbers of both confirmed new cases and confirmed deaths are unreliable, but on the other hand, no other data are available. Some models, e.g. IHME 2020, are capable of estimating the true number of infections, but this involves making a number of additional assumptions, and is based partly on the reporting of testing-dependent data. Also, the relation between the true number of infections and the number of deaths has not been well studied to date and requires a number of assumptions. Using the number of infections seems to be the easiest way of obtaining basic data on the COVID-19 infection dynamics in a given country, so long as one is aware that publicly shared data show the number of confirmed cases and not the number of real infections and takes this into account when interpreting the results.

In conclusion, the method based on fitting Johnson CDF curves to the cumulative number of confirmed cases is straightforward, well-known and easy to use. It yields curves that are extremely well fitted to the data, and the basic parameters of COVID-19 infection dynamics thus obtained are easy to interpret and use in further statistical analyses by researchers from fields other than epidemiology, e.g. sociology, biology and ecology. While deepening our understanding of the COVID-19 pandemic, it may also be useful for making short-term predictions, although caution is advisable in such cases.

## Data Availability

The data used in this study was obtained from Our World in Data COVID-19 dataset and are available at https://ourworldindata.org/coronavirus

## Acknowledgements

Both authors made equal contributions to this study and are listed alphabetically. This study was financed by the statutory funds of the Institute of Nature Conservation, Polish Academy of Sciences, and the Faculty of Applied Mathematics, AGH University of Science and Technology. We thank Magdalena Lenda and Piotr Skórka for their useful comments and suggestions.

