## SupplementaryMaterials for "A new, simple method of describing the COVID-19 trajectory and dynamics in any country based on Johnson Cumulative Distribution Function fitting"

**Supplementary Materials**

**Fitted Johnson Cumulative Distribution Functions**

Johnson Cumulative Distribution Functions fitted to the data obtained for 80 countries on six continents are shown in Figs. S1-S6 (Africa, Fig. S1; Asia, Fig. S2; Europe, Fig. S3; North America, Fig. S4; Oceania, Fig. S5; South America, Fig. S6). The formulas and R^2^ values of each fitted curve are listed in Table S1.

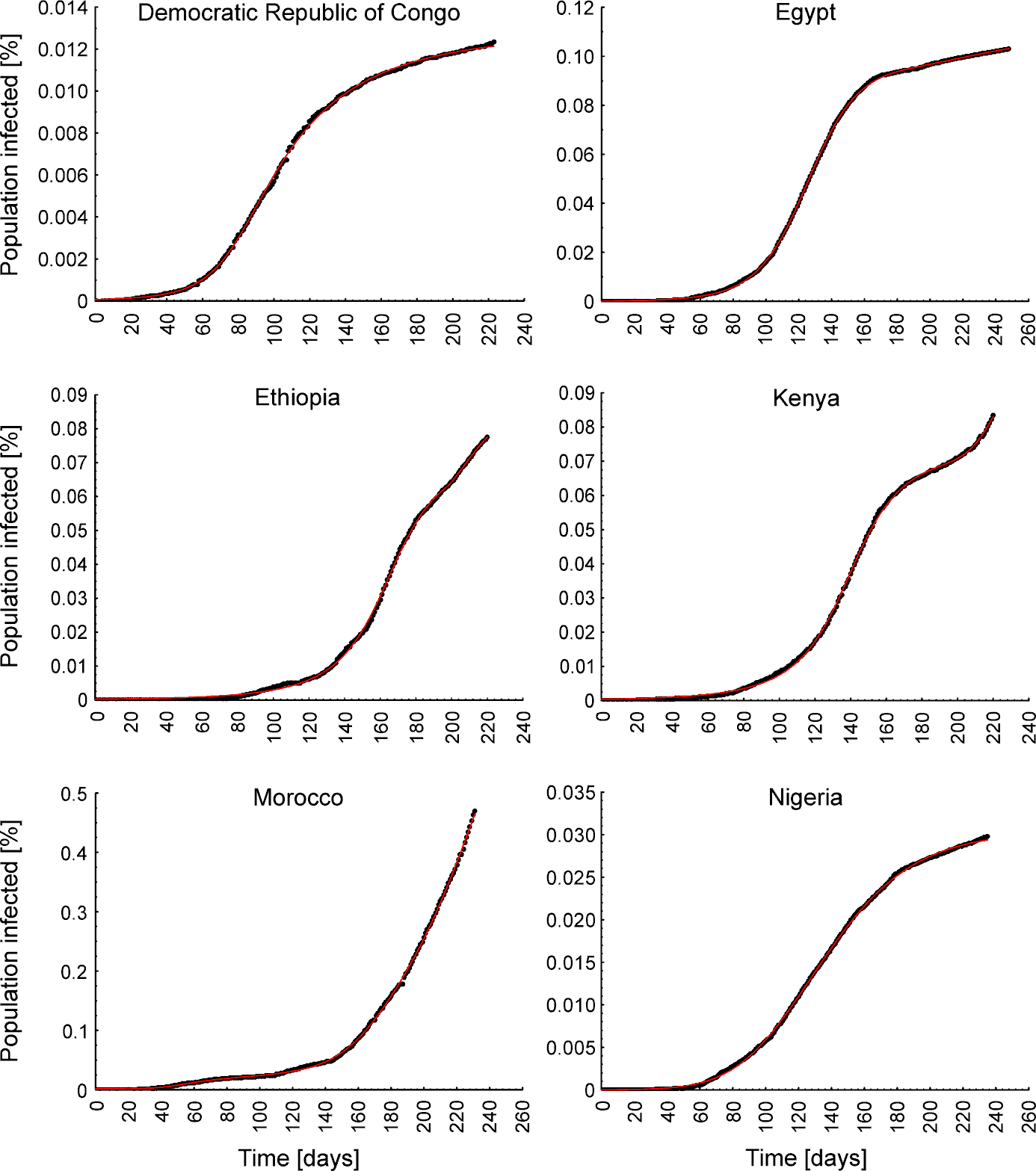

Figure S1. Johnson Cumulative Distribution Functions (red lines) fitted to the data on COVID-19 trajectories in African countries.

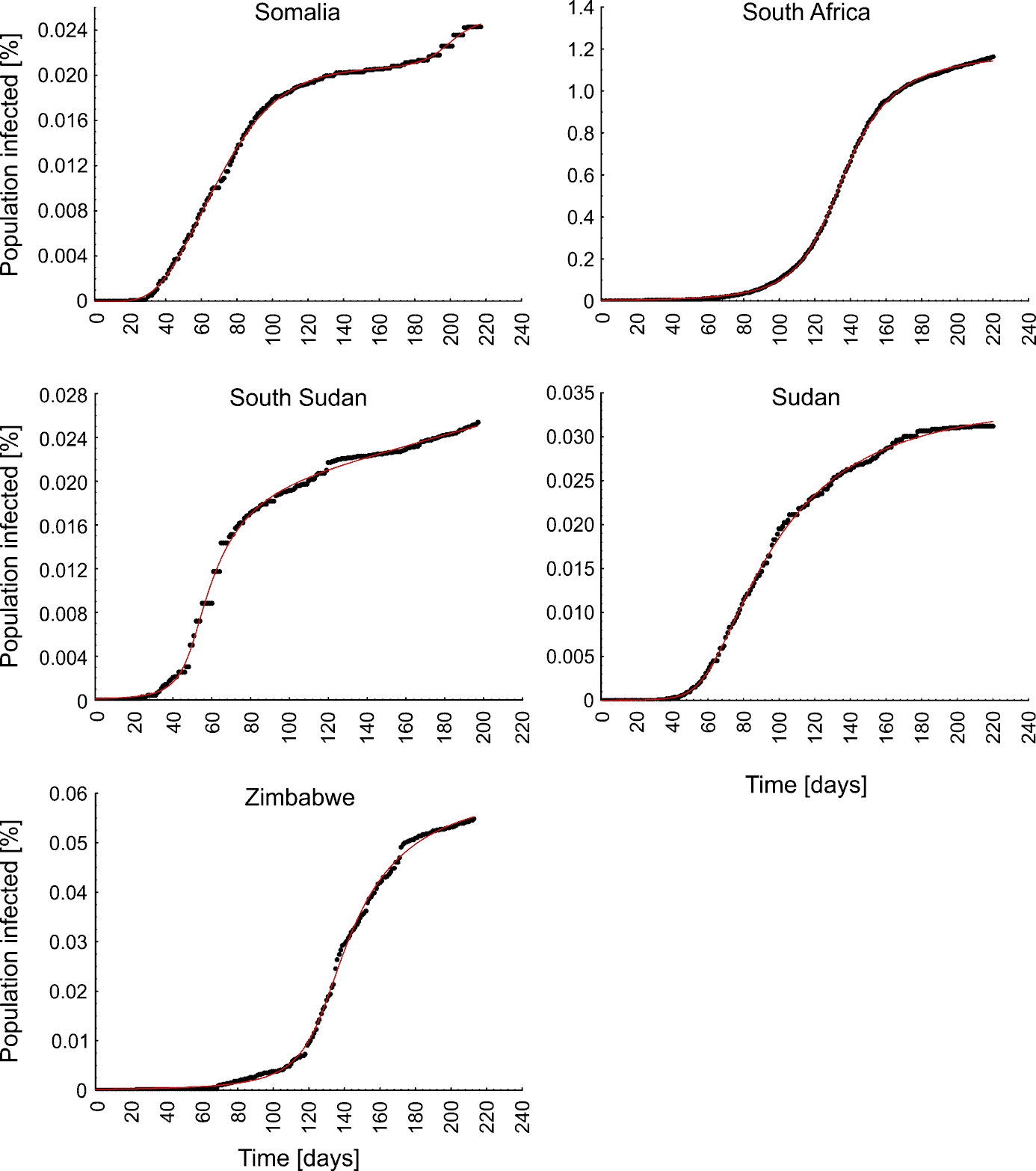

Figure S1 continued. Johnson Cumulative Distribution Functions (red lines) fitted to the data on COVID-19 trajectories in African countries.

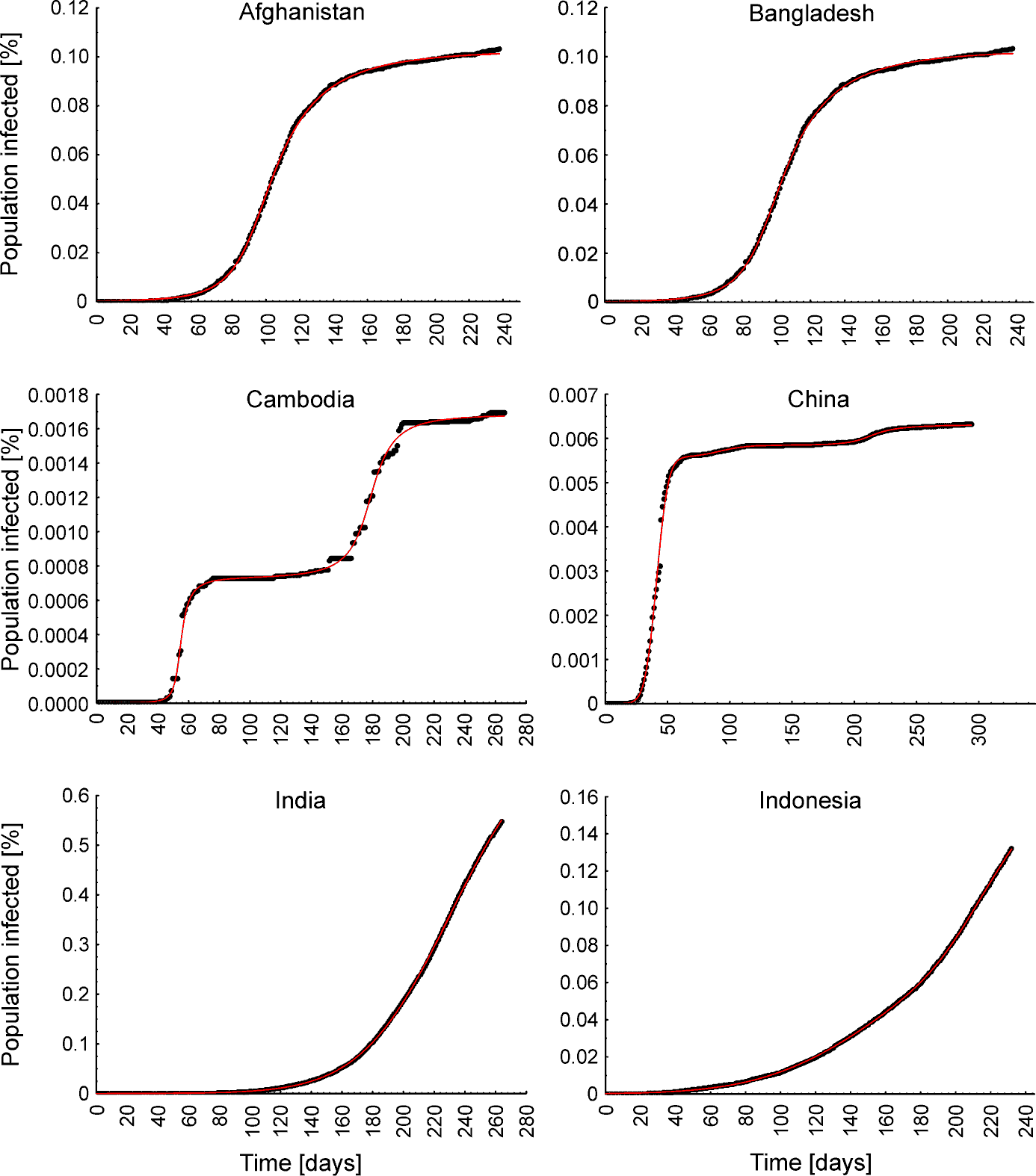

Figure S2. Johnson Cumulative Distribution Functions (red lines) fitted to the data on COVID-19 trajectories in Asian countries.

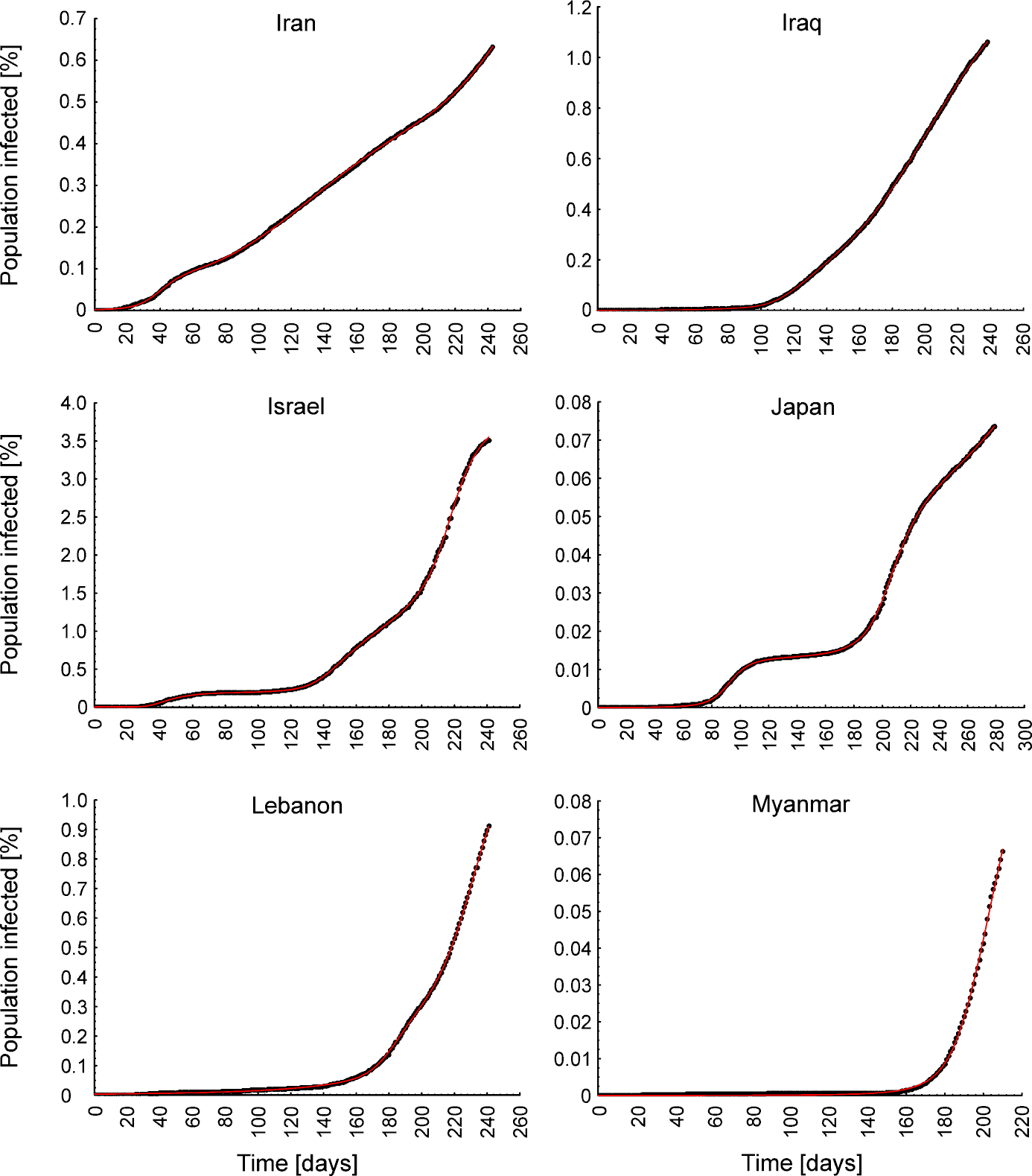

Figure S2 continued. Johnson Cumulative Distribution Functions (red lines) fitted to the data on COVID-19 trajectories in Asian countries.

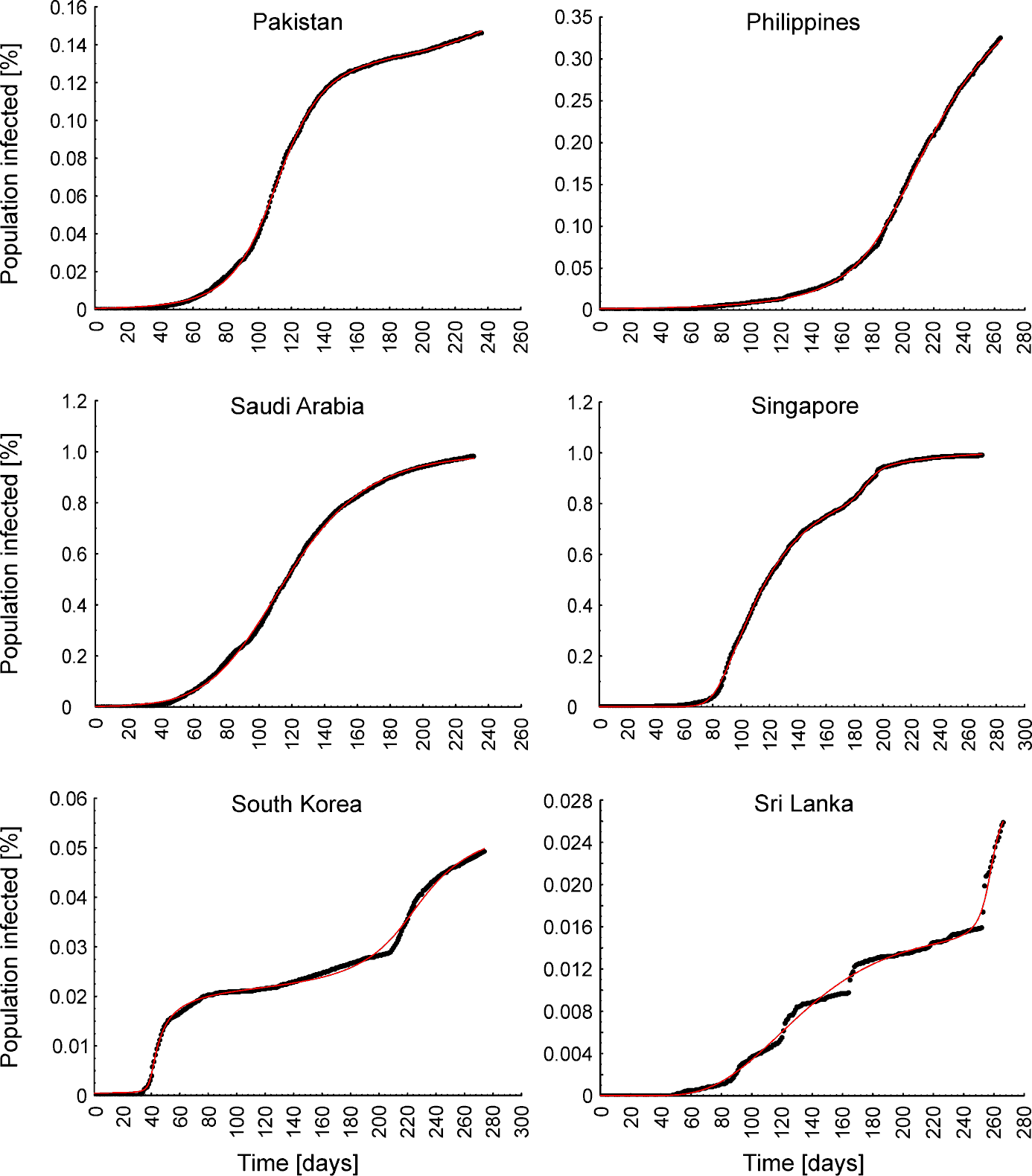

Figure S2 continued. Johnson Cumulative Distribution Functions (red lines) fitted to the data on COVID-19 trajectories in Asian countries.

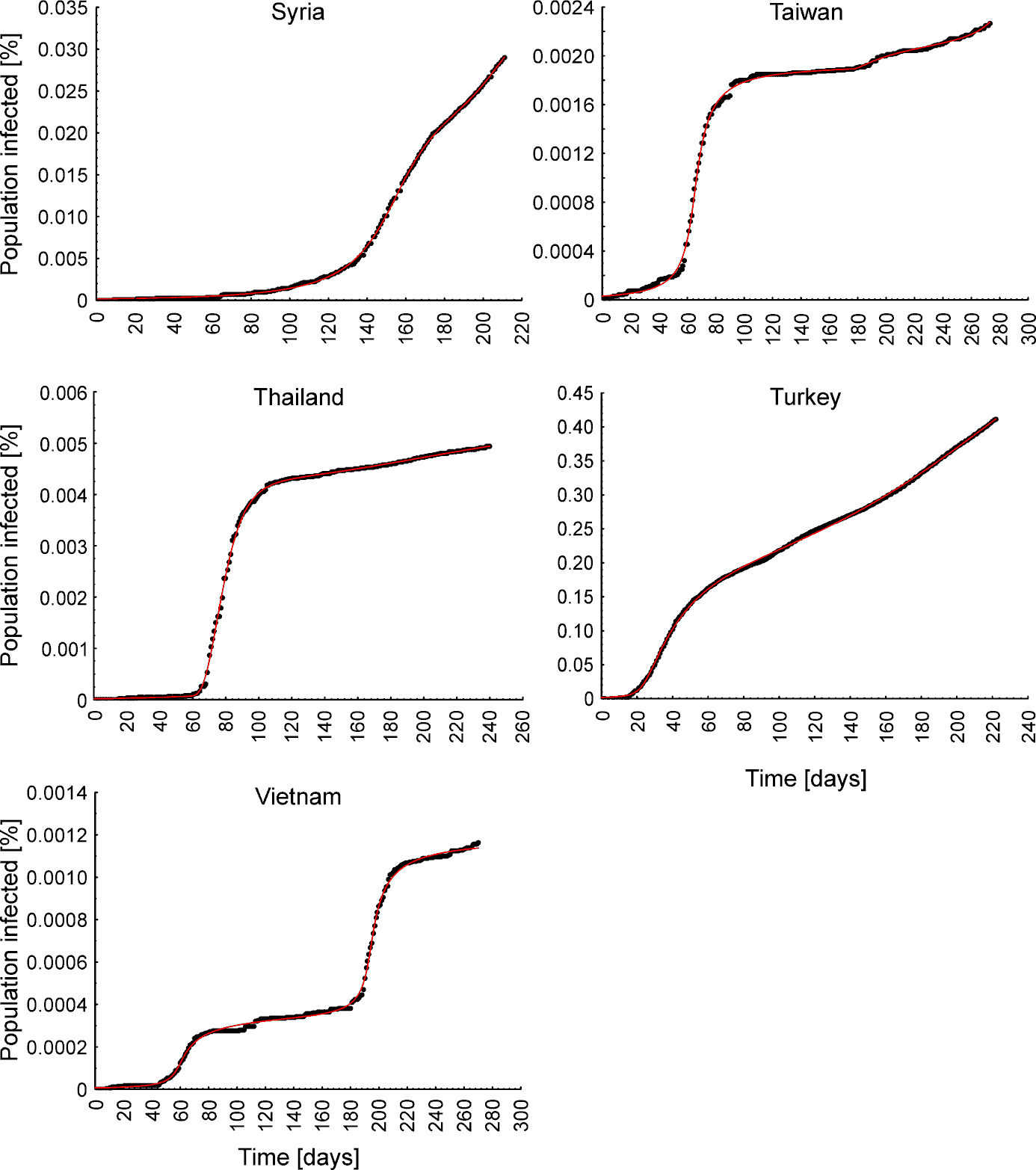

Figure S2 continued. Johnson Cumulative Distribution Functions (red lines) fitted to the data on COVID-19 trajectories in Asian countries.

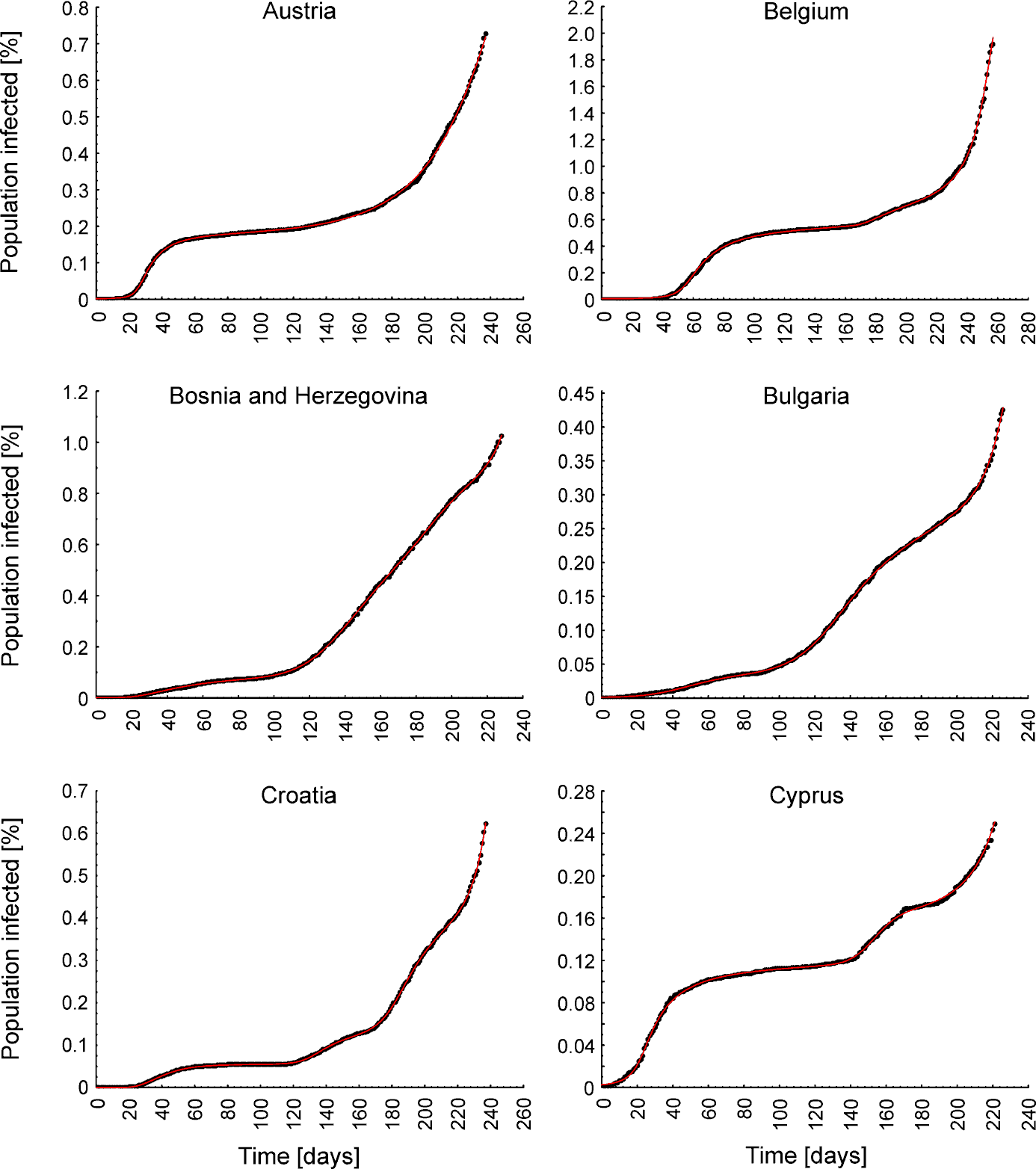

Figure S3. Johnson Cumulative Distribution Functions (red lines) fitted to the data on COVID-19 trajectories in European countries.

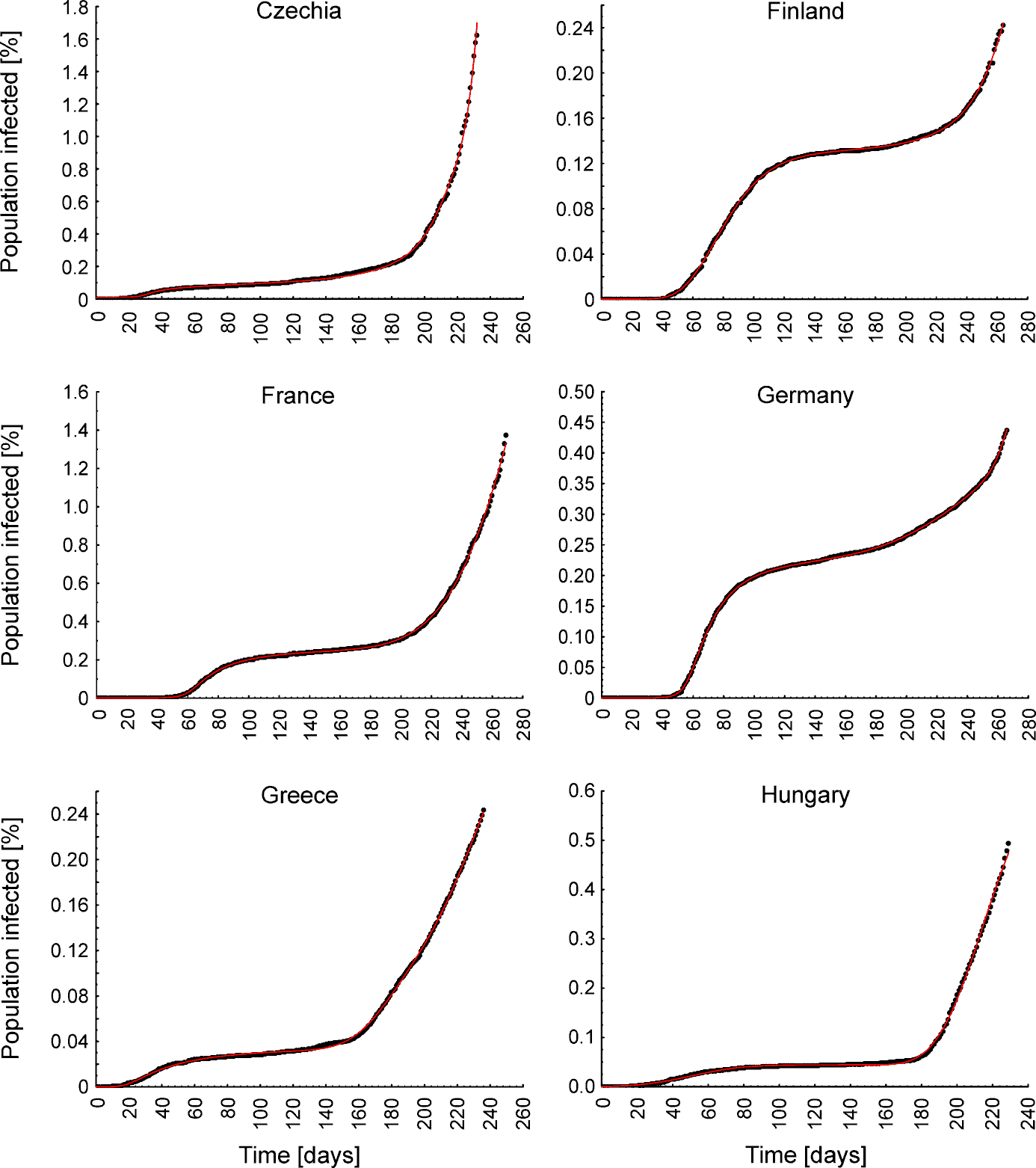

Figure S3 continued. Johnson Cumulative Distribution Functions (red lines) fitted to the data on COVID-19 trajectories in European countries.

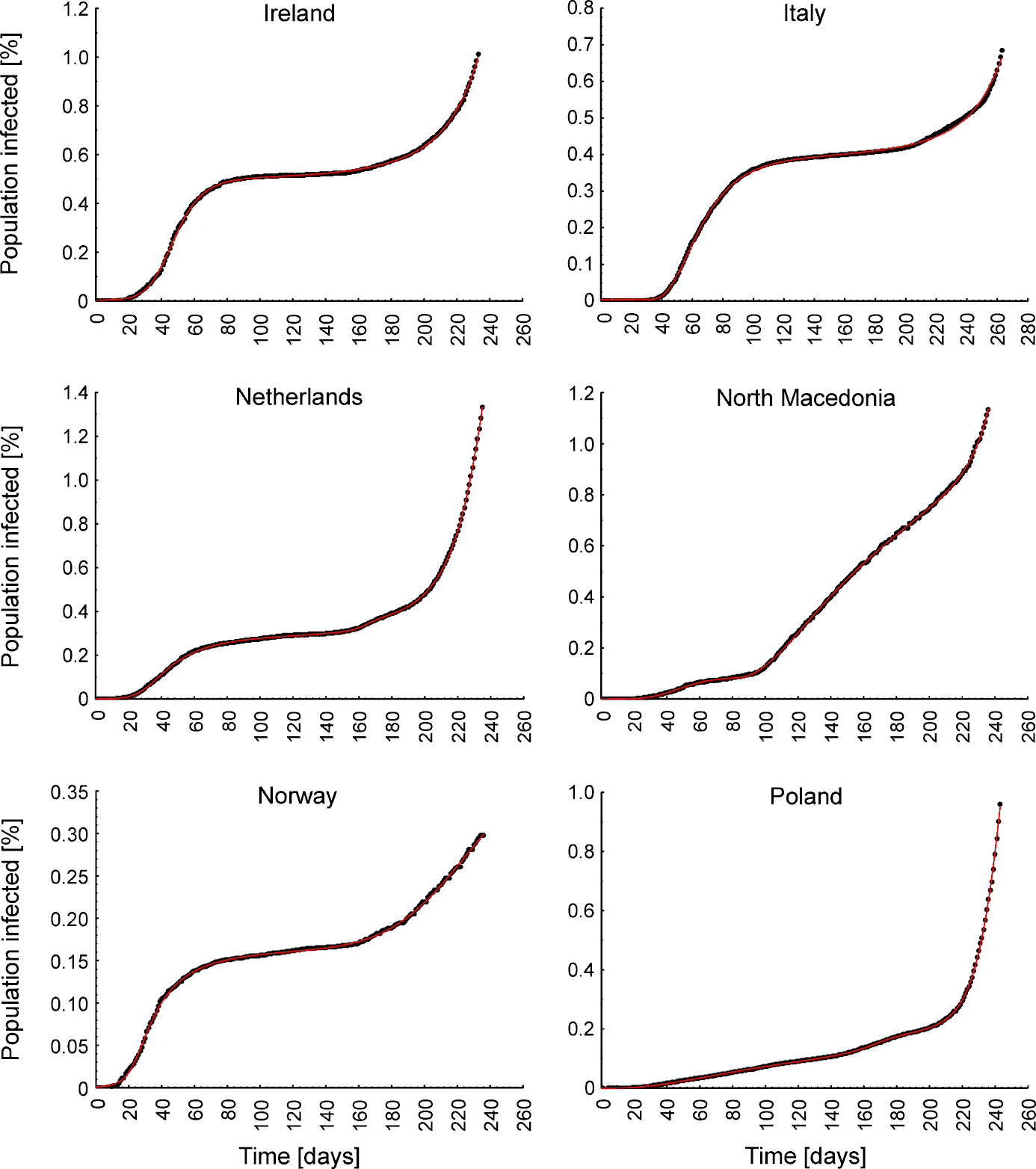

Figure S3 continued. Johnson Cumulative Distribution Functions (red lines) fitted to the data on COVID-19 trajectories in European countries.

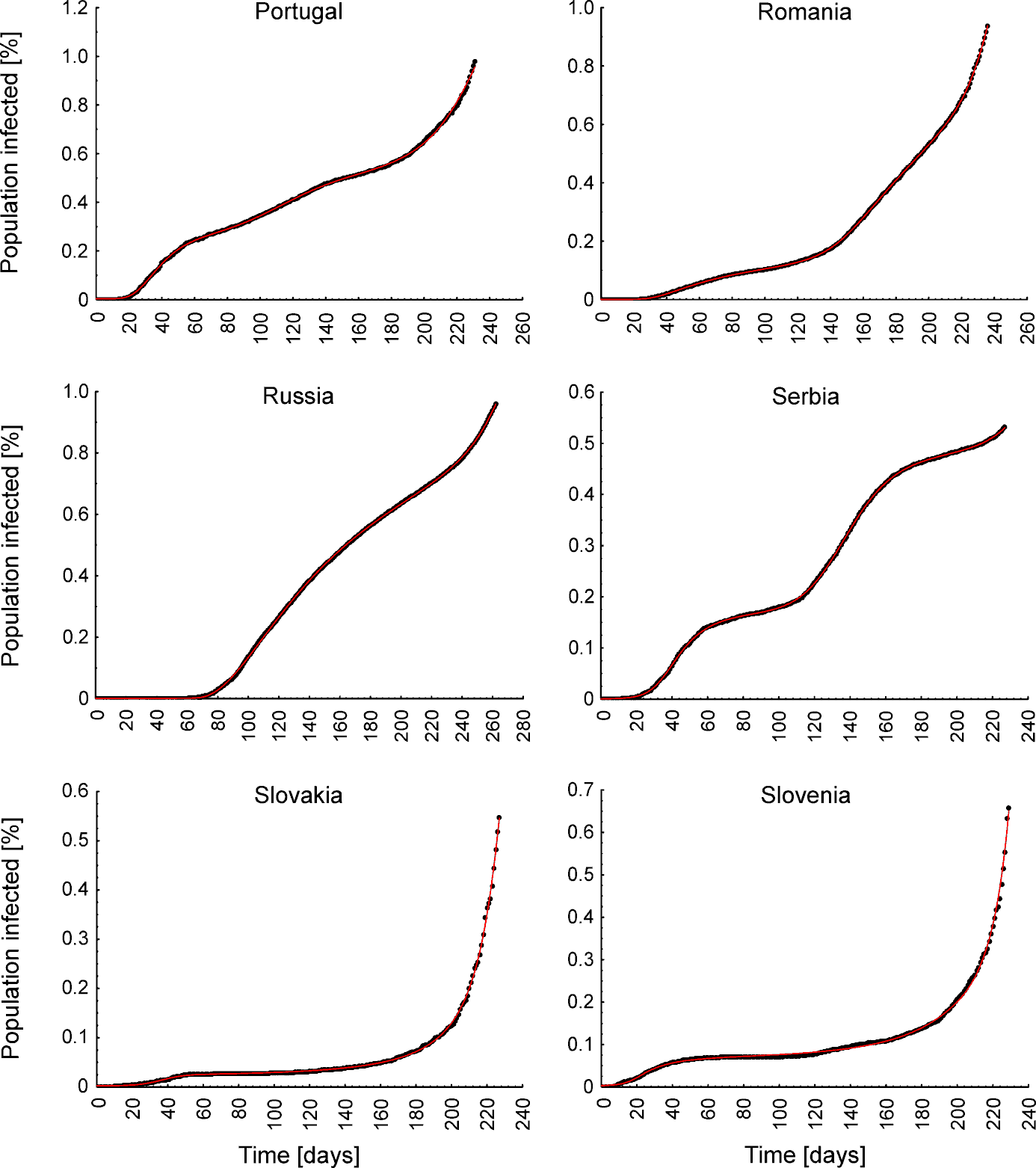

Figure S3 continued. Johnson Cumulative Distribution Functions (red lines) fitted to the data on COVID-19 trajectories in European countries.

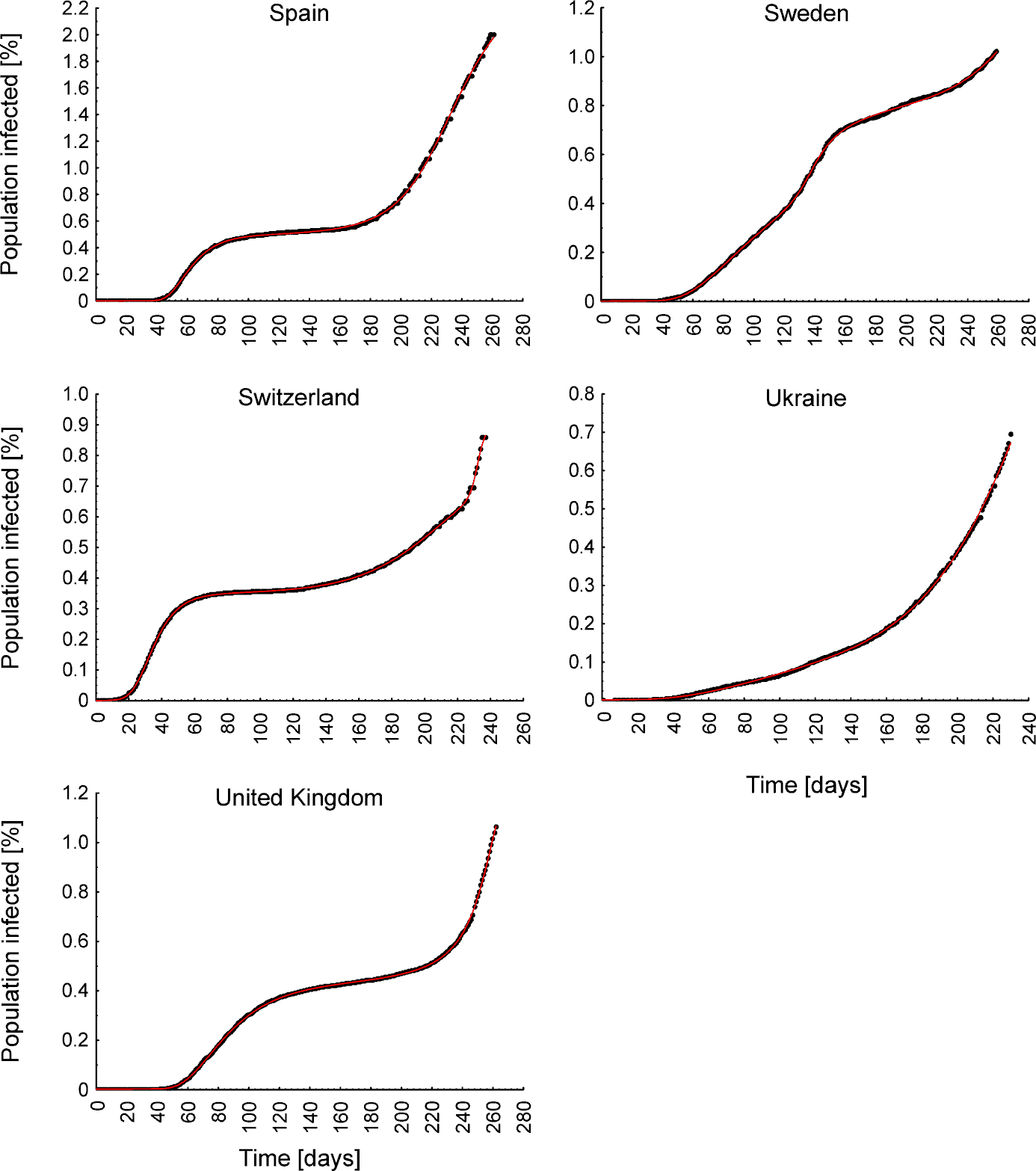

Figure S3 continued. Johnson Cumulative Distribution Functions (red lines) fitted to the data on COVID-19 trajectories in European countries.

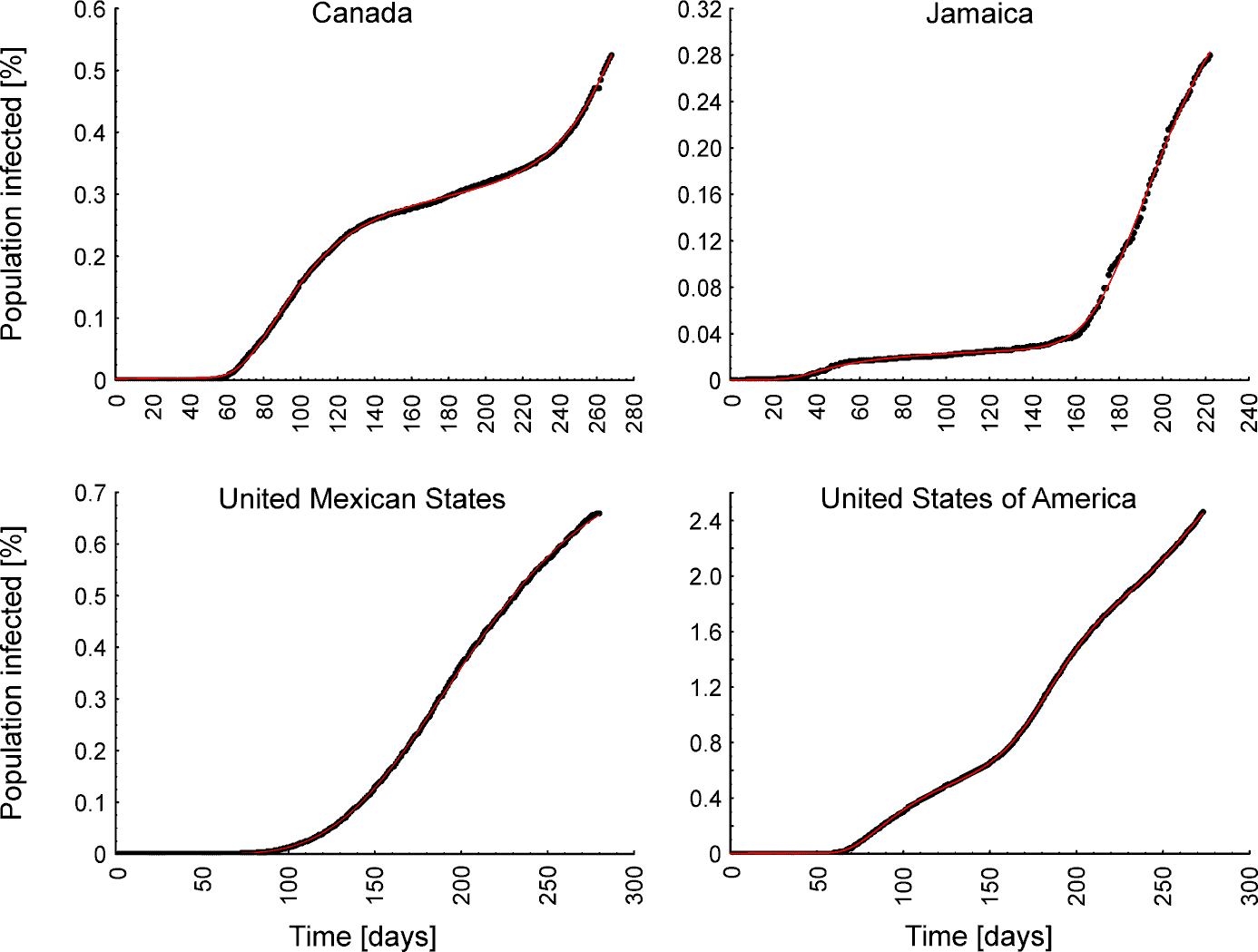

Figure S4. Johnson Cumulative Distribution Functions (red lines) fitted to the data on COVID-19 trajectories in North American countries.

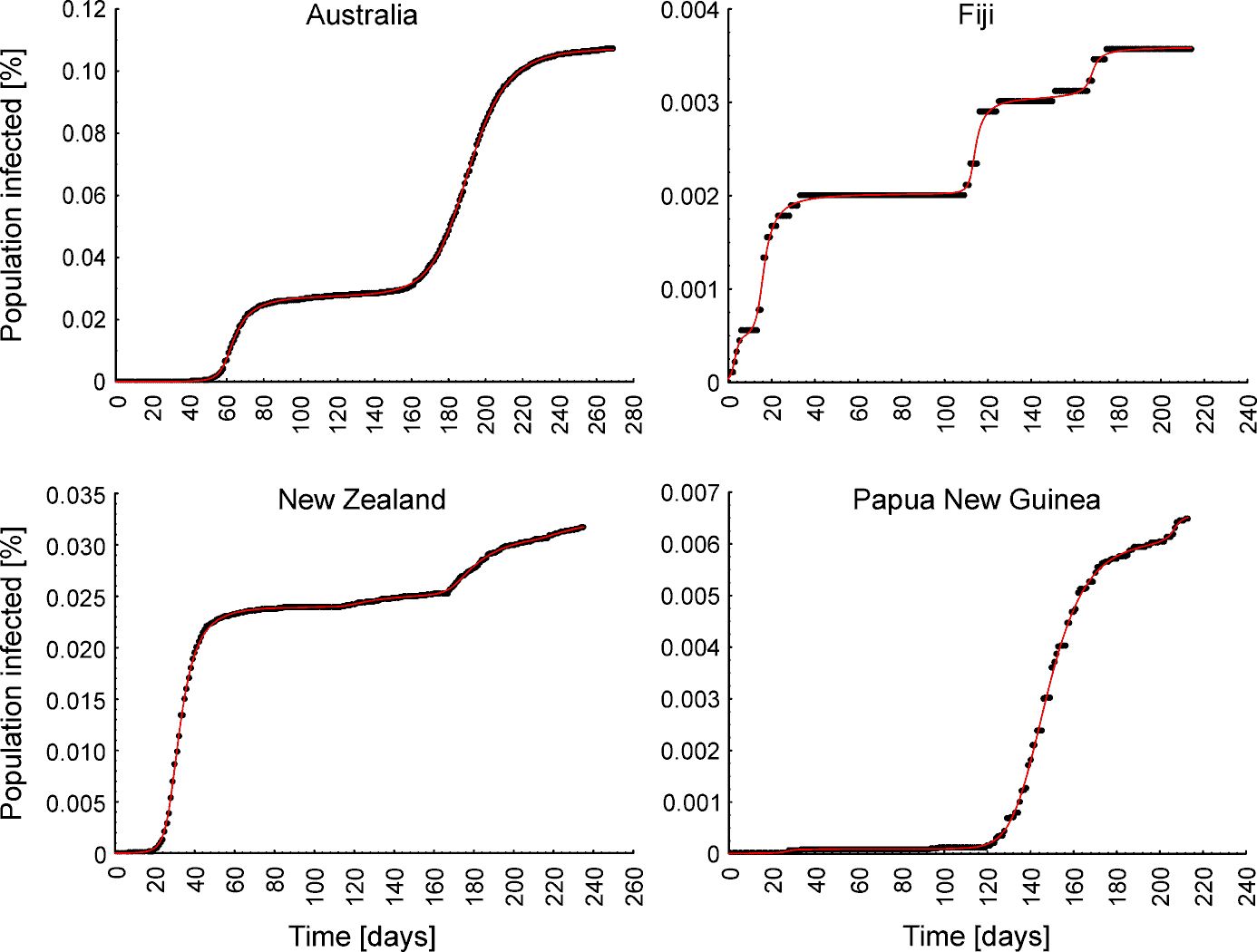

Figure S5. Johnson Cumulative Distribution Functions (red lines) fitted to the data on COVID-19 trajectories in countries in Oceania.

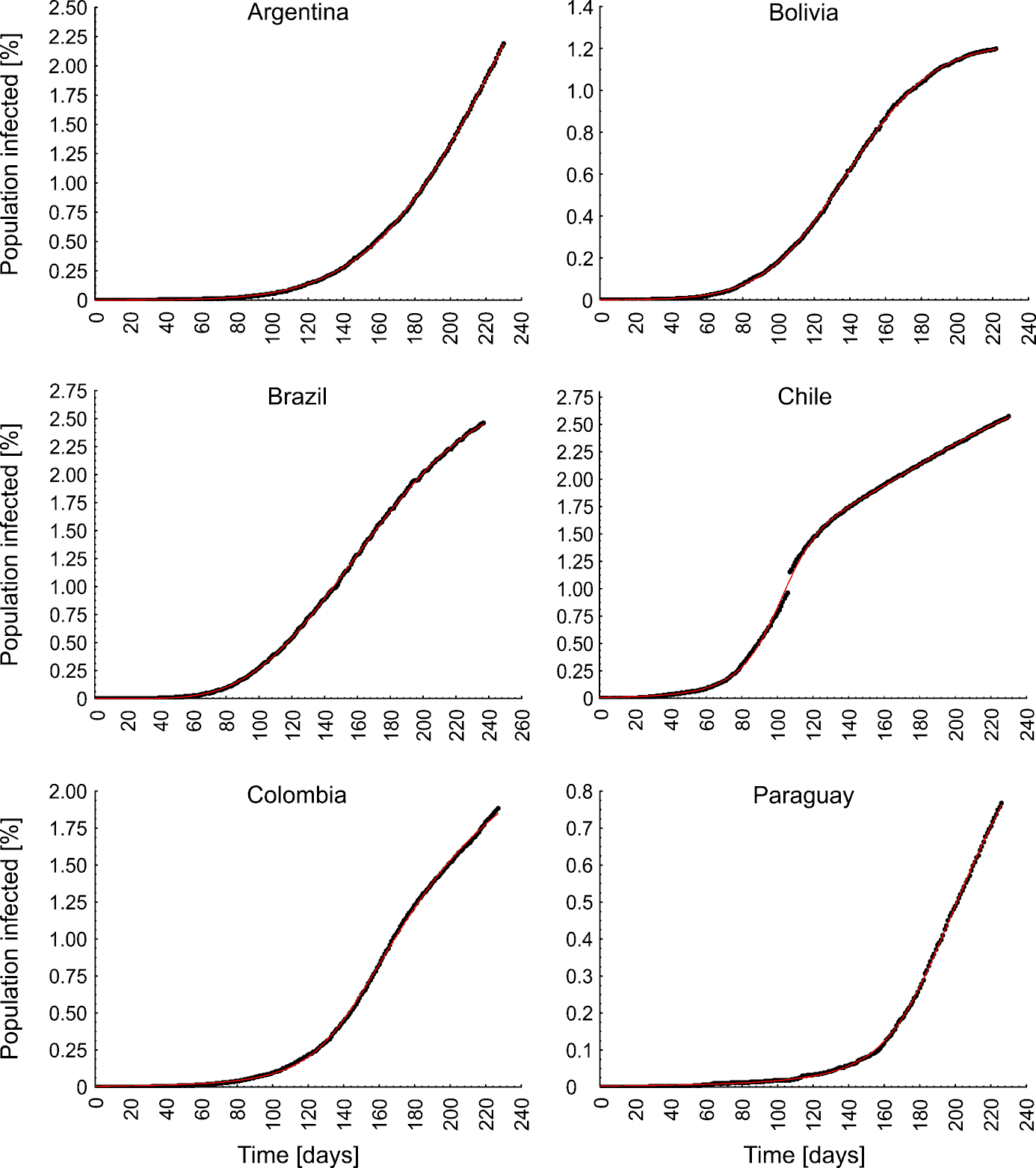

Figure S6. Johnson Cumulative Distribution Functions (red lines) fitted to the data on COVID-19 trajectories in South American countries.

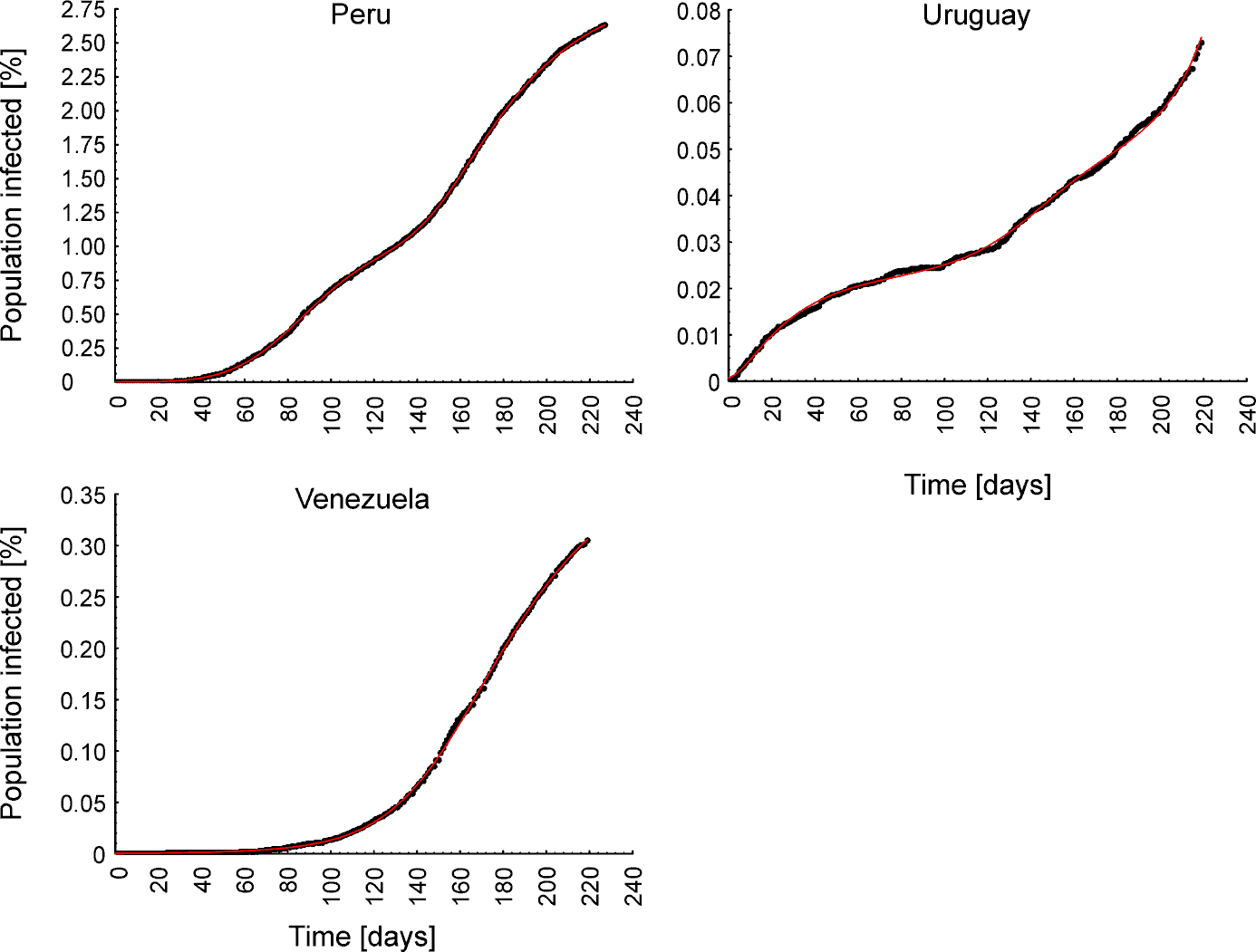

Figure S6 continued. Johnson Cumulative Distribution Functions (red lines) fitted to the data on COVID-19 trajectories in South American countries.

Table S1. Number of infection waves (*N_w_*) and fitted Johnson Cumulative Distribution Functions with R^2^ for each of 80 countries.

| **Region** | **Country** | **N_w_** | **Fitted curve** | **R^2^** |
| --- | --- | --- | --- | --- |
| Africa | Democratic Republic of Congo | 1 | *W(t)=0.012907*F_118.898,70.7757,4.41941,62.2782_(t)* | 0.99970 |
| Africa | Egypt | 2 | *W(t)=0.0972531*F_125.352,30.6693,0.00050384,3.06174_(t)+0.0122929*F_260.741,54.8242,1.6964,5.5283_(t)* | 0.99992 |
| Africa | Ethiopia | 2 | *W(t)=0.0724824*F_164.787,40.9673,0.000152085,22.0159_(t)+0.0382048*F_298.465,166.884,9.18943,308.199_(t)* | 0.99949 |
| Africa | Kenya | 2 | *W(t)=0.0731944*F_139.094,39.836,0.00513622,17.513_(t)+0.0474776*F_233.749,18.9369,0.53122,0.515896_(t)* | 0.99970 |
| Africa | Morocco | 3 | *W(t)=0.0141559*F_59.9864,17.0538,1.02118,1.91984_(t)+0.0834356*F_171.999,20.6423,0.00891419,0.000862216_(t)*  *+0.805248*F_241.998,67.234,1.46249,15.7144_(t)* | 0.99970 |
| Africa | Nigeria | 1 | *W(t)=0.0303193*F_138.608,46.3461,0.618287,1.23014_(t)* | 0.99981 |
| Africa | Somalia | 2 | *W(t)=0.0205073*F_71.4445,26.9654,0.669545,0.386906_(t)+0.00498195*F_202.499,28.8392,1.87492,50.8692_(t)* | 0.99936 |
| Africa | South Africa | 1 | *W(t)=1.17782*F_139.383,39.416,1.9068,30.4609_(t)* | 0.99982 |
| Africa | South Sudan | 2 | *W(t)=0.0215857*F_72.4221,51.5002,10.3282,487.988_(t)+0.00710007*F_187.292,68.6273,0.000721799,0.0014819_(t)* | 0.99832 |
| Africa | Sudan | 1 | *W(t)=0.0340264*F_115.566,73.978,4.41345,50.7955_(t)* | 0.99935 |
| Africa | Zimbabwe | 1 | *W(t)=0.0619585*F_159.383,81.6095,10.5104,581.313_(t)* | 0.99892 |
| Asia | Afghanistan | 1 | *W(t)=0.102767*F_111.081,41.5161,3.53541,51.35_(t)* | 0.99980 |
| Asia | Bangladesh | 1 | *W(t)=0.348623*F_232.815,234.097,6.96886,154.172_(t)* | 0.99987 |
| Asia | Cambodia | 2 | *W(t)=0.000728299*F_56.3424,9.91615,5.24715,196.964_(t)+0.000953623*F_179.33,27.6622,0.900536,120.952_(t)* | 0.99873 |
| Asia | China | 3 | *W(t)=0.00557997*F_41.168,7.71032,0.00789602,0.335735_(t)+0.000256916*F_92.3289,16.2606,0.108518,0.0104642_(t)+0.000482547*F_219.72,49.6171,6.06916,438.392_(t)* | 0.99922 |
| Asia | India | 1 | *W(t)=0.722943*F_230.67,49.6579,0.00668056,0.674791_(t)* | 0.99991 |
| Asia | Indonesia | 2 | *W(t)=0.0762435*F_153.723,54.2348,0.00615382,0.282283_(t)+0.124304*F_246.064,63.3083,2.65265,18.7225_(t)* | 0.99993 |
| Asia | Iran | 3 | *W(t)=0.0639611*F_39.748,10.2852,0.00678327,0.000767145_(t)+0.373225*F_128.03,49.4976,0.00801498,0.00690689_(t)+0.539529*F_257.097,48.9543,0.379968,4.75668_(t)* | 0.99989 |

Table S1 continued. Number of infection waves (*N_w_*) and fitted Johnson Cumulative Distribution Functions with R^2^ for each of 80 countries.

| **Region** | **Country** | **N_w_** | **Fitted curve** | **R^2^** |
| --- | --- | --- | --- | --- |
| Asia | Iraq | 2 | *W(t)=0.131308*F_129.153,17.7527,0.70030,0.894485_(t)+1.43748*F_225.835,71.2414,1.4831,6.34081_(t)* | 0.99997 |
| Asia | Israel | 3 | *W(t)=0.178083*F_47.7947,12.8397,1.23695,2.84856_(t)+1.07353*F_175.363,53.3451,4.36542,53.2556_(t)+2.75224*F_217.455,36.3955,0.00702233,106.2_(t)* | 0.99984 |
| Asia | Japan | 3 | *W(t)=0.0129294*F_92.9975,17.4181,0.367292,11.0077_(t)+0.0549041*F_223.087,55.3649,6.65743,198.157_(t)+0.024011*F_283.843,31.084,3.54246e-005,2.01303_(t)* | 0.99994 |
| Asia | Lebanon | 2 | *W(t)=0.125332*F_182.461,11.1118,0.00612455,0.00375655_(t)+1.22259*F_231.754,51.8948,0.170241,72.7_(t)* | 0.99983 |
| Asia | Myanmar | 1 | *W(t)=0.231405*F_278.06,158.573,9.57165,353.868_(t)* | 0.99929 |
| Asia | Pakistan | 2 | *W(t)=0.134688*F_112.052,32.9648,0.292952,8.18107_(t)+0.0498235*F_263.816,43.7466,0.0064386,0.00248769_(t)* | 0.99978 |
| Asia | Philippines | 1 | *W(t)=0.464499*F_258.656,148.513,7.92314,264.871_(t)* | 0.99954 |
| Asia | Saudi Arabia | 1 | *W(t)=0.992046*F_120.187,44.7246,0.754774,2.17753_(t)* | 0.99954 |
| Asia | Singapore | 2 | *W(t)=0.904572*F_127.87,52.8599,3.45234,28.0657_(t)+0.111026*F_187.985,8.46479,0.00217685,0.00510362_(t)* | 0.99986 |
| Asia | South Korea | 2 | *W(t)=0.0207541*F_54.585,36.5873,18.056,1952.21_(t)+0.0355237*F_233.456,110.451,4.31021,550.873_(t)* | 0.99710 |
| Asia | Sri Lanka | 2 | *W(t)=0.0151098*F_137.562,47.6004,0.766996,0.792379_(t)+0.0133252*F_258.358,13.7617,0.555389,62.4503_(t)* | 0.99570 |
| Asia | Syria | 2 | *W(t)=0.0240074*F_154.728,45.707,0.000259146,89.2068_(t)+0.024922*F_295.176,230.068,17.5589,1806.64_(t)* | 0.99975 |
| Asia | Taiwan | 3 | *W(t)=0.00190445*F_68.5568,44.8968,15.0248,9536.08_(t)+0.000229525*F_242.32,114.578,10.3972,426.548_(t)+0.00198782*F_302.701,52.1451,11.127,1434.31_(t)* | 0.99931 |
| Asia | Thailand | 2 | *W(t)=0.00410741*F_80.1918,11.1578,1.21911,2.76417_(t)+0.00121732*F_198.408,88.4168,0.00801526,0.327718_(t)* | 0.99971 |
| Asia | Turkey | 2 | *W(t)=0.218575*F_62.5161,68.5548,9.28242,325.306_(t)+0.44534*F_236.638,92.6851,0.149186,0.0249466_(t)* | 0.99964 |
| Asia | Vietnam | 2 | *W(t)=0.000353952*F_89.8007,188.62,141.486,549249_(t)+0.00083114*F_205.597,92.0535,98.4422,286635_(t)* | 0.99917 |

Table S1 continued. Number of infection waves (*N_w_*) and fitted Johnson Cumulative Distribution Functions with R^2^ for each of 80 countries.

| **Region** | **Country** | **N_w_** | **Fitted curve** | **R^2^** |
| --- | --- | --- | --- | --- |
| Europe | Austria | 2 | *W(t)=0.17527*F_37.1483,18.2457,4.85582,72.0865_(t)+1.72405*F_258.79,56.6247,0.331707,6.14541_(t)* | 0.99952 |
| Europe | Belgium | 3 | *W(t)=0.530032*F_75.9429,39.6471,7.05439,180.883_(t)+2.30285*F_254.694,46.6561,0.00106393,1480.24_(t)+0.130713*F_194.409,23.6728,2.51733,13.0585_(t)* | 0.99947 |
| Europe | Bosnia and Herzegovina | 4 | *W(t)=0.0739929*F_53.246,28.8839,2.0354,8.1794_(t)+0.538101*F_157.025,36.0771,1.3177,4.50388_(t)+0.309098*F_199.417,73.828,6.7710,557.55_(t)+1.99763*F_250.332,38.156,12.058,967.95_(t)* | 0.99987 |
| Europe | Bulgaria | 3 | *W(t)=0.0269585*F_47.5719,17.7046,0.00146355,0.00307107_(t)+0.249875*F_156.12,61.25,3.92436,53.962_(t)+0.372563*F_227.127,41.3025,0.00119748,1039.35_(t)* | 0.99985 |
| Europe | Croatia | 4 | *W(t)=0.0541405*F_42.3028,14.5852,1.2054,2.7003_(t)+0.0699166*F_142.426,19.0722,2.0881,12.541_(t)+0.265581*F_195.313,26.1445,2.4423,20.489_(t)+ 1.84424*F_254.091,25.2158,3.8395,84.862_(t)* | 0.99987 |
| Europe | Cyprus | 3 | *W(t)=0.115092*F_38.8547,41.5975,11.51,674.693_(t)+0.0427542*F_153.321,10.3041,0.00302284,0.00496755_(t)+0.685592*F_241.113,30.4744,0.166811,83.0336_(t)* | 0.99941 |
| Europe | Czechia | 3 | *W(t)=0.0740236*F_53.422,57.53,15.7487,1335.66_(t)+0.580889*F_207.855,84.1427,0.00343905,5776.05_(t)+3.10353*F_239.091,46.3418,17.1092,6298.29_(t)* | 0.99871 |
| Europe | Finland | 2 | *W(t)=0.129189*F_83.5325,23.8704,0.810138,1.19927_(t)+0.343111*F_277.954,46.5977,1.54627,37.4286_(t)* | 0.99976 |
| Europe | France | 2 | *W(t)=0.261132*F_90.6015,51.8933,7.89034,221.255_(t)+4.10448*F_297.808,44.9799,0.00799113,0.0101135_(t)* | 0.99946 |
| Europe | Germany | 3 | *W(t)=0.222966*F_76.4158,26.2769,3.50858,31.082_(t)+0.181065*F_233.356,52.5644,5.64595e-006,1.93187_(t)+0.20839*F_274.96,29.9089,5.70031,159.122_(t)* | 0.99993 |
| Europe | Greece | 3 | *W(t)=0.0348153*F_61.9783,63.5106,6.17117,112.763_(t)+0.103529*F_181.986,20.5377,0.00202955,0.00197367_(t)*  *+0.319133*F_258.944,42.0616,1.28874,3.10161_(t)* | 0.99963 |
| Europe | Hungary | 2 | *W(t)=0.0441469*F_50.6497,22.4631,0.0013507,0.00260687_(t)+0.627712*F_219.048,23.3921,0.37232,0.258861_(t)* | 0.99897 |

Table S1 continued. Number of infection waves (*N_w_*) and fitted Johnson Cumulative Distribution Functions with R^2^ for each of 80 countries.

| **Region** | **Country** | **N_w_** | **Fitted curve** | **R^2^** |
| --- | --- | --- | --- | --- |
| Europe | Ireland | 2 | *W(t)=0.497913*F_48.895,15.585,0.699044,2.31044_(t)+2.83079*F_257.14,40.6004,0.195159,34.0572_(t)* | 0.99966 |
| Europe | Italy | 2 | *W(t)=0.384347*F_69.7684,22.9268,1.59687,4.86194_(t)+0.731665*F_270.257,66.8497,0.00872201,404.655_(t)* | 0.99940 |
| Europe | Netherlands | 3 | *W(t)=0.287691*F_51.6405,28.4184,2.80814,19.3507_(t)+0.0365955*F_164.368,6.55801,0.00694339,0.00650985_(t)+3.1847*F_248.207,36.8726,1.49748,24.4172_(t)* | 0.99991 |
| Europe | North Macedonia | 3 | *W(t)=0.0829176*F_48.9238,15.1939,0.275836,0.0210793_(t)+1.04243*F_220.912,154.087,4.53121,51.4201_(t)+1.51616*F_247.943,25.1638,2.7862e-005,54.5253_(t)* | 0.99988 |
| Europe | Norway | 2 | *W(t)=0.163703*F_42.5197,32.1053,5.21729,85.2685_(t)+0.26025*F_238.661,52.5603,0.848722,2.75276_(t)* | 0.99959 |
| Europe | Poland | 3 | *W(t)=0.132827*F_102.361,55.4639,0.6934,0.0223507_(t)+0.0668263*F_166.381,24.9569,0.00170481,11.9708_(t)+2.34471*F_250.565,18.5969,0.0915285,2.19396_(t)* | 0.99981 |
| Europe | Portugal | 3 | *W(t)=0.258048*F_42.5502,17.561,1.63762,5.12868_(t)+0.215328*F_112.182,32.6685,0.00328498,1.71916_(t)+1.03118*F_233.218,58.2843,0.00419217,65.8505_(t)* | 0.99961 |
| Europe | Romania | 3 | *W(t)=0.133803*F_89.3277,70.5765,4.3139,46.2637_(t)+0.780209*F_239.423,29.0504,0.0001,42.0172_(t)+0.783166*F_268.079,225.04,9.86942,384.941_(t)* | 0.99996 |
| Europe | Russia | 2 | *W(t)=0.824778*F_175.61,102.354,3.27165,23.6967_(t)+1.02793*F_281.37,42.8491,1e-005,25.5494_(t)* | 0.99995 |
| Europe | Serbia | 3 | *W(t)=0.170125*F_47.0028,19.5063,1.85867,9.94486_(t)+0.30192*F_139.757,23.7944,0.442784,1.54262_(t)+1.07463*F_264.903,30.5913,1.51098,28.8457_(t)* | 0.99995 |
| Europe | Slovakia | 2 | *W(t)=0.0210139*F_36.623,9.83823,0.00649583,0.00114204_(t)+1.44706*F_237.886,53.2076,8.2407,872.307_(t)* | 0.99913 |
| Europe | Slovenia | 3 | *W(t)=0.0667705*F_31.0458,22.1511,3.82183,44.5024_(t)+0.0372809*F_153.216,45.378,3.13201,21.426_(t)+3.10237*F_240.203,26.5982,0.00343161,356.051_(t)* | 0.99779 |
| Europe | Spain | 2 | *W(t)=0.488605*F_64.7353,15.3781,1.33086,3.32056_(t)+2.13144*F_247.923,61.8614,2.724,31.7884_(t)* | 0.99955 |

Table S1 continued. Number of infection waves (*N_w_*) and fitted Johnson Cumulative Distribution Functions with R^2^ for each of 80 countries.

| **Region** | **Country** | **N_w_** | **Fitted curve** | **R^2^** |
| --- | --- | --- | --- | --- |
| Europe | Sweden | 3 | *W(t)=0.489846*F_108.244,45.1005,1.56794,4.67754_(t)+0.297079*F_141.705,25.0865,2.42091,46.5798_(t)+0.814077*F_279.847,54.6232,0.00622152,10.7975_(t)* | 0.99987 |
| Europe | Switzerland | 3 | *W(t)=0.347761*F_36.8963,13.1397,1.09462,2.6412_(t)+0.45431*F_209.667,50.124,0.0052421,3.03597_(t)+0.20404*F_232.143,4.56782,0.00999514,3.2024_(t)* | 0.99971 |
| Europe | Ukraine | 2 | *W(t)=0.0974848*F_101.361,42.2545,0.591188,0.0564594_(t)+2.53384*F_293.439,91.9928,1.15615,4.51605_(t)* | 0.99958 |
| Europe | United Kingdom | 2 | *W(t)=0.424517*F_91.7018,30.9686,1.67723,5.51722_(t)+1.11024*F_258.58,45.4118,3.7409e-005,286.879_(t)* | 0.99986 |
| North America | Canada | 2 | *W(t)=0.280839*F_104.241,32.1361,1.5151,4.35129_(t)+0.517431*F_270.257,68.205,0.001574,94.8645_(t)* | 0.99971 |
| North America | Jamaica | 2 | *W(t)=0.0293465*F_89.9085,126.443,14.6287,1088.16_(t)+0.39252*F_217.746,51.5602,2.46008,14.1724_(t)* | 0.99913 |
| North America | Mexico | 1 | *W(t)=0.744036*F_206.989,59.3047,0.429441,0.0393411_(t)* | 0.99987 |
| North America | United States of America | 3 | *W(t)=0.550603*F_103.596,30.6493,1.66289,5.29818_(t)+1.19572*F_185.159,30.4121,0.156334,1.74146_(t)+1.84218*F_312.212,113.282,4.70041,76.1051_(t)* | 0.99997 |
| Oceania | Australia | 2 | *W(t)=0.17527*F_37.1483,18.2457,4.85582,72.0865_(t)+1.72405*F_258.79,56.6247,0.331707,6.14541_(t)* | 0.99995 |
| Oceania | Fiji | 3 | *W(t)=0.00202481*F_14.5684,13.9534,0.000356559,76.5454_(t)+0.00101567*F_115.978,7.33466,6.06082,148.182_(t)+0.000546777*F_168.152,20.8587,0.000183706,19851.4_(t)* | 0.99497 |
| Oceania | New Zealand | 4 | *W(t)=0.0239738*F_34.354,10.3045,2.70678,22.131_(t)+0.00130462*F_141.203,39.8374,6.6160,139.061_(t)+0.00603697*F_189.027,29.2594,4.9602,72.4903_(t)+0.00156923*F_244.737,35.1765,3.403,25.9784_(t)* | 0.99982 |
| Oceania | Papua New Guinea | 3 | *W(t)=9.57574e05*F_27.1931,90.834,0.00514512,1.18301e+007_(t)+0.00591296*F_149.554,16.4096,0.98963,2.91694_(t)+ 0.000620503*F_206.608,33.8333,0.0014881,108048_(t)* | 0.99949 |

Table S1 continued. Number of infection waves (*N_w_*) and fitted Johnson Cumulative Distribution Functions with R^2^ for each of 80 countries.

| **Region** | **Country** | **N_w_** | **Fitted curve** | **R^2^** |
| --- | --- | --- | --- | --- |
| South America | Argentina | 2 | W(t)=0.17527*F_37.1483,18.2457,4.85582,72.0865_*(t)*+1.72405*F_258.79,56.6247,0.331707,6.14541_*(t)* | 0.99992 |
| South America | Bolivia | 1 | W(t)=1.22183*F_139.642,39.0547,0.0962302,0.317568_*(t)* | 0.99992 |
| South America | Brazil | 1 | W(t)=2.74199*F_167.255,53.0097,0.284768,0.236245_*(t)* | 0.99992 |
| South America | Chile | 2 | W(t)=1.81944*F_102.247,28.2296,0.0048419,8.30444_*(t)*+1.30239*F_236.372,83.3318,1.9275,7.26942_*(t)* | 0.99966 |
| South America | Colombia | 1 | W(t)=2.79391*F_232.005,160.021,6.71011,152.059_*(t)* | 0.99971 |
| South America | Paraguay | 1 | W(t)=1.92033*F_351.719,398.677,15.815,1351.45_*(t)* | 0.99979 |
| South America | Peru | 2 | W(t)=0.933518*F_86.145,25.0466,0.00462452,0.00837006_*(t)*+2.06124*F_190.799,55.982,2.64435,17.9234_*(t)* | 0.99995 |
| South America | Uruguay | 3 | W(t)=0.0240813*F_37.6414,38.1737,3.22348,22.897_*(t)*+0.0214958*F_146.316,28.0122,0.144591,0.00416435_*(t)*+0.109209*F_238.163,66.7847,3.37775,455.887_*(t)* | 0.99855 |
| South America | Venezuela | 1 | W(t)=0.398944*F_188.667,61.1926,1.55804,8.43544_*(t)* | 0.99978 |

**Fitting Johnson curves to the ongoing wave; forecasting possibilities**

The accuracy of forecasts is discussed on the basis of data relating to the first wave of infections in the United Kingdom. The U.K. was selected because it is both highly populated (67886004) and, among European countries, has carried out the largest number of tests: a mean 126.33 of tests per 1000 since the beginning of the pandemic up to 19 November 2020.

The first wave of infections was described using the Johnson distribution curve, and parameters *P_inf_* , *Q_2.5%_*, *M*, *Q_97.5%_* , *T_i_* , *T_d_* and *T* were calculated. The values obtained for *Q_2.5%_*, M and *Q_97.5%_* indicated that the first wave of infections in the U.K. started on the 51^st^ and finished on the 170^th^ day of the epidemic, while the wave peaked on the 74^th^ day. Then, a series of forecasts was made by fitting Johnson distribution curves to the data cut to the first 60, 70, 80, 90, 100, 110, 120, 130, 140, 150 and 160 days of the epidemic. *P_inf_* ,*Q_2.5%_*, *M*, *Q_97.5%_*, *T_i_* , *T_d_* and *T* were calculated for each forecast: the results are listed in Table S2. In addition, the percentage difference between the actual and predicted percentage of infections on each day was calculated for each forecast (starting from the forecast day to its end at t=170; see Figure S7). Figure S8 illustrates example forecasts based on days 60, 90, 210 and 150 compared to the Johnson distribution curve fitted to the complete data.

The results showed that the predicted parameters describing the day of the wave peak, the day of the end of the infection wave, as well as the durations of the wave and its increase and decrease, did not differ much from the values obtained using the complete dataset for the first wave of infections in the U.K. (Table S2). However, the percentages of the population infected predicted using only days 70 and 80 were ca 0.1% lower than the actual figures. This means that, if recalculated to the number of infections, the predicted number of infections was ca 68000 lower than the actual number of infections. This suggests that such early predictions (prior to the peak) made using Johnson CDF fitting should focus on predicting the day of the peak rather than the number of infections. Also, for early forecasts, the predicted daily percentages of infections differed from the actual percentages of infections by more than 10% in the longer term (Fig. S7). On the other hand, the predictions made after the wave had peaked were consistent with observations.

It also needs to be highlighted that this curve fitting method was designed primarily not for making forecasts but for obtaining easily interpretable parameters describing the past trajectory of COVID-19 infections. Thus, extreme caution is advisable when forecasting the future trajectory of the infection wave and its parameters (see the Discussion).

Table S2. Parameters describing the percentage of the infected population (*P_inf_*), the day of the start of the infection wave (*Q_2.5%_*), the day when the wave peaked (*M*), the day of the end of the infection wave (*Q_97.5%_*), the duration of the wave increase (*T_i_*), the duration of the wave decrease (*T_d_*) and the duration of the wave of infections (*T*) for each forecast using a different number of days from the beginning of the epidemic (t=0) to the end of the first infection wave (t=170).

| Days used | *P_inf_* | *Q_2.5%_* | *M* | *Q_97.5%_* | *T_i_* | *T_d_* | *T* |
| --- | --- | --- | --- | --- | --- | --- | --- |
| 60 (9 days after wave start) | 0.434241 | 51 | 74 | 157 | 23 | 83 | 106 |
| 70 | 0.334370 | 49 | 69 | 165 | 20 | 96 | 116 |
| 80 | 0.338205 | 49 | 69 | 169 | 20 | 100 | 120 |
| 90 | 0.414673 | 50 | 75 | 161 | 25 | 86 | 111 |
| 100 | 0.444636 | 51 | 75 | 170 | 24 | 95 | 119 |
| 110 | 0.459856 | 51 | 75 | 180 | 24 | 105 | 129 |
| 120 | 0.444350 | 51 | 75 | 170 | 24 | 95 | 120 |
| 130 | 0.433287 | 50 | 75 | 163 | 25 | 88 | 112 |
| 140 | 0.430665 | 50 | 76 | 161 | 26 | 85 | 111 |
| 150 | 0.433254 | 50 | 75 | 163 | 25 | 88 | 113 |
| 160 | 0.437829 | 50 | 75 | 168 | 25 | 93 | 118 |
| 170 (complete wave) | 0.424517 | 52 | 74 | 170 | 22 | 96 | 118 |

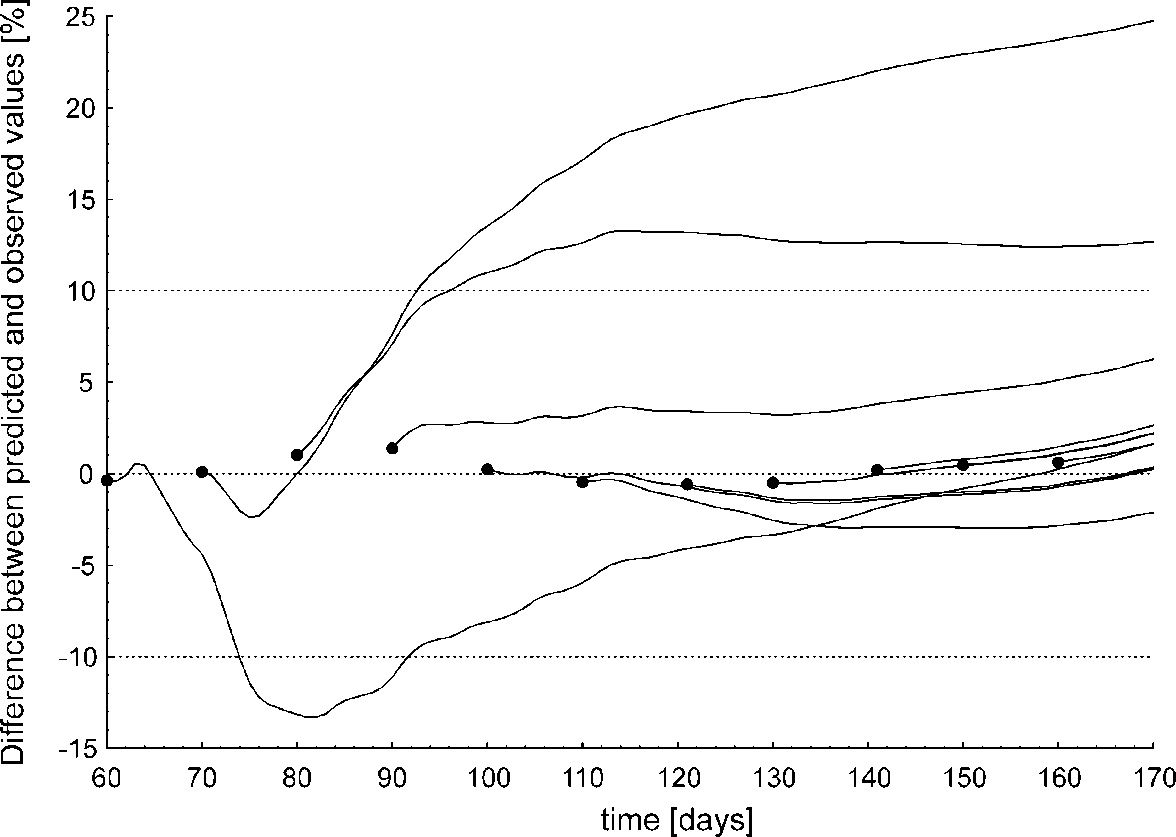

Fig.S7. The percentage difference between the actual percentage of the population infected and the predicted percentage of the population infected for 11 forecasts using different numbers of days (60, 70, 80, 90, 100, 110, 120, 130, 140, 150 and 160), starting from the forecast day to the end at t=170.

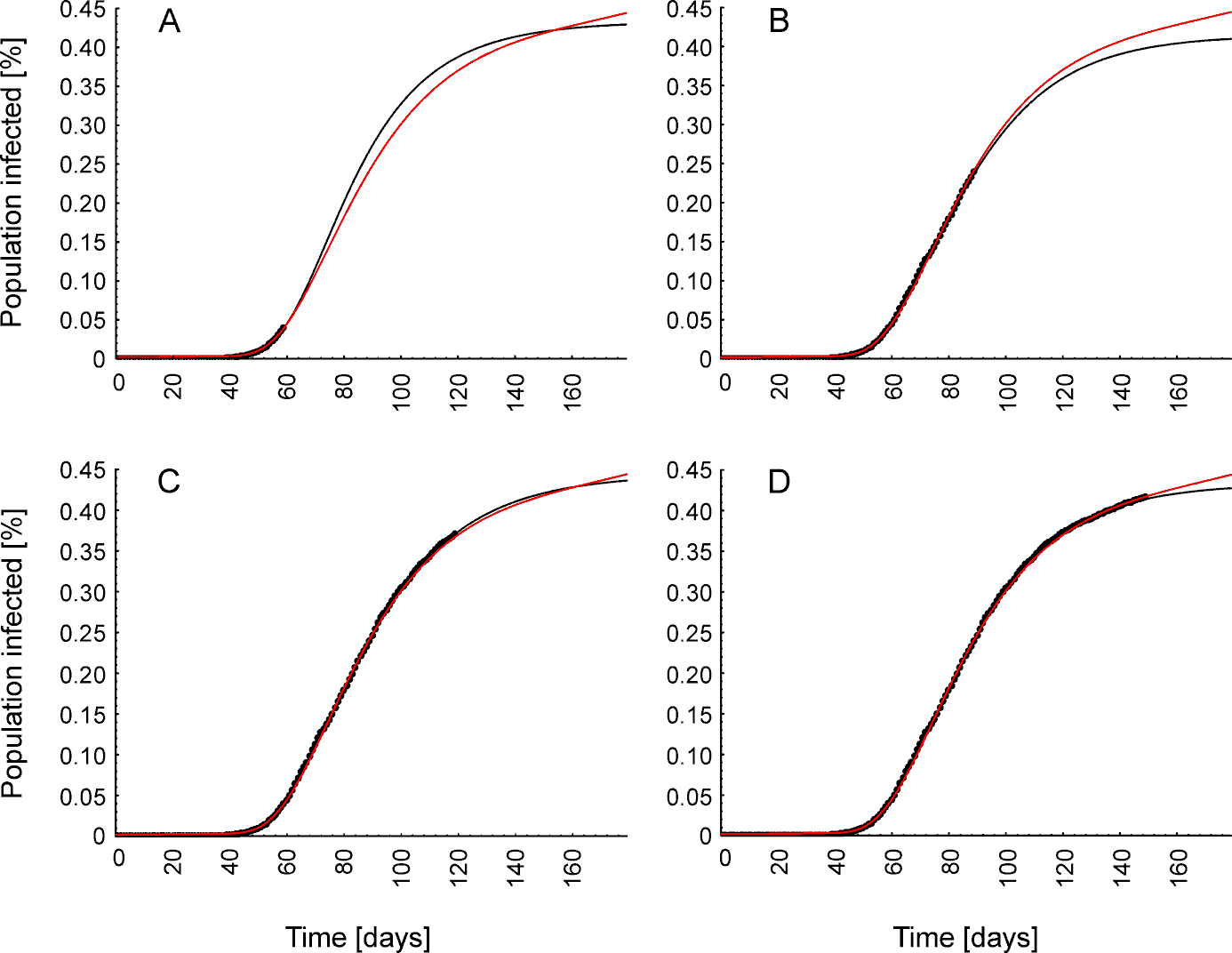

Fig. S8. Examples of predictions of the future trajectory of the infection wave (black line) using data (black dots) from days 60 (A), 90 (B), 120 (C) and 150 (D) in comparison to the actual trajectory of the infection wave (red line).
